# Preventive small-quantity lipid-based nutrient supplements reduce severe wasting and severe stunting among young children: an individual participant data meta-analysis of randomized controlled trials

**DOI:** 10.1101/2022.06.16.22276521

**Authors:** Kathryn G. Dewey, Charles D. Arnold, K. Ryan Wessells, Elizabeth L. Prado, Souheila Abbeddou, Seth Adu-Afarwuah, Hasmot Ali, Benjamin F. Arnold, Per Ashorn, Ulla Ashorn, Sania Ashraf, Elodie Becquey, Kenneth H. Brown, Parul Christian, John M. Colford, Sherlie J. L. Dulience, Lia C.H. Fernald, Emanuela Galasso, Lotta Hallamaa, Sonja Y. Hess, Jean H. Humphrey, Lieven Huybregts, Lora L. Iannotti, Kaniz Jannat, Anna Lartey, Agnes Le Port, Jef L. Leroy, Stephen P. Luby, Kenneth Maleta, Susana L. Matias, Mduduzi NN Mbuya, Malay K. Mridha, Minyanga Nkhoma, Clair Null, Rina R. Paul, Harriet Okronipa, Jean-Bosco Ouédraogo, Amy J. Pickering, Andrew J. Prendergast, Marie Ruel, Saijuddin Shaikh, Ann M. Weber, Patricia Wolff, Amanda Zongrone, Christine P. Stewart

## Abstract

**Background:** Meta-analyses show that small-quantity lipid-based nutrient supplements (SQ-LNS) reduce child wasting and stunting. There is little information regarding effects on severe wasting or stunting.

**Objective:** We aimed to identify the effect of SQ-LNS on severe wasting (weight-for-length z-score < −3) and severe stunting (length-for-age z-score < −3).

**Methods:** We conducted a two-stage meta-analysis of individual participant data from 14 randomized controlled trials of SQ-LNS provided to children 6 to 24 mo of age. We generated study-specific and subgroup estimates of SQ-LNS vs. control and pooled the estimates using fixed-effects models. We used random effects meta-regression to examine study-level effect modifiers. In sensitivity analyses, we examined whether results differed depending on study arm inclusion criteria and types of comparisons.

**Results:** Q-LNS provision led to a relative reduction of 31% in severe wasting (Prevalence Ratio, PR 0.69 (0.55, 0.86), n=34,373) and 17% in severe stunting (PR 0.83 (95% CI: 0.78, 0.90), n=36,795). Results were similar in most of the sensitivity analyses but somewhat attenuated when comparisons using passive control arms were excluded: PR 0.74 (0.57, 0.96), n=26,327 for severe wasting and PR 0.88 (0.81, 0.95), n=28,742 for severe stunting. Study-level characteristics generally did not significantly modify the effects of SQ-LNS, but results suggested greater effects of SQ-LNS in sites with greater burdens of wasting or stunting, or with poorer water quality or sanitation.

**Conclusions:** Including SQ-LNS in preventive interventions to promote healthy child growth and development is likely to reduce rates of severe wasting and stunting. Registered at www.crd.york.ac.uk/PROSPERO as CRD42019146592.

## Introduction

The global prevalence of stunting (length-for-age z-score (LAZ) < −2) among children under 5 years of age was estimated to be 22% in 2021 (1), which represents 149 million children. Severe stunting (LAZ < −3) likely affects 40-50% of that total (2). For wasting (weight-for-length z-score (WLZ) < −2), the estimated cross-sectional prevalence was 6.7% in 2021 (45.4 million), but that is an underestimate of the total annual burden of wasting because children often cycle in and out of being wasted due to seasonal and other factors. The total annual burden of wasting may be 3-6 times greater than an estimate based on cross-sectional prevalence, depending on the country and context (3, 4). In a pooled analysis of 21 longitudinal cohorts under 2 y of age (the most vulnerable period for wasting (5, 6)), 6.5% of children were wasted at a specific visit but 29.2% experienced at least one episode of wasting by 24 mo of age (7). The global prevalence of severe wasting (WLZ < −3) was 2% in 2020 (13.6 million in 2021), but again, this is an underestimate of the total burden which may be as much as 7-10 times higher (4). Risk of mortality is 5.5 times higher among children with severe stunting and 11.6 times higher among children with severe wasting, compared to children with z-scores > −1 for LAZ or WLZ, respectively (8). Moreover, severe malnutrition in early life is associated with serious adverse consequences for subsequent health and development (9, 10).

There has been inadequate progress in reducing rates of stunting and wasting, both moderate and severe (1), and in recent years rates of child malnutrition have been rising in areas affected by armed conflict, climate change and the economic disruptions brought about by the COVID-19 pandemic (6, 11). Thus, there is a pressing need to identify strategies to reduce severe undernutrition among young children. Recent initiatives such as the Global Action Plan on Child Wasting (12) and the development of guidelines for the prevention and treatment of wasting in infants and children (13) reflect the growing awareness of the urgent need for evidence-based actions.

Although the etiology of severe stunting and wasting is complex and multi-factorial (14–17), inadequate dietary intake plays a pivotal role. During the complementary feeding period from 6 to 24 mo of age, diets often lack adequate amounts of nutrients that are critical for growth (18), in part because of the high cost of nutrient-rich foods for low-income families. Fortified products, such as fortified blended foods and products used for home fortification including micronutrient powders (MNP) and small-quantity lipid-based nutrient supplements (SQ-LNS) (18), can help fill these nutrient gaps. SQ-LNS provide multiple micronutrients embedded in a small amount of food (∼110-120 kcal/d) that also provides energy, protein, and essential fatty acids (19). SQ-LNS were designed for the prevention of undernutrition, whereas larger quantities of LNS are generally aimed at treatment of moderate and severe wasting. While there have been numerous intervention trials to evaluate treatments for severe wasting, there is very little evidence regarding interventions that are effective for prevention of this life-threatening condition (20, 21).

In a recent individual participant data (IPD) analysis of 14 randomized controlled trials, we found a 12-14% lower prevalence of stunting, wasting and underweight, as well as reductions in developmental delay, anemia and micronutrient deficiencies among children who received SQ-LNS during the complementary feeding period (22–25). We did not include severe wasting or severe stunting in that set of analyses because we already had a large list of outcomes to examine, and also because a key objective of that work was examining individual-level effect modification, which is problematic for rare outcomes such as severe wasting. However, a previous Cochrane review and meta-analysis of LNS (including both SQ-LNS and medium-quantity LNS) (26) reported both of these outcomes. The authors reported a 15% reduction in severe stunting (RR 0.85, 95% CI 0.74 to 0.98) based on 5 studies (6,151 participants); they did not find an effect on severe wasting but only 3 studies (2,329 participants) included this outcome. Given the strengthened global commitment to combatting severe malnutrition, there has been interest in updating the findings for these two outcomes using the much larger IPD dataset. Therefore, the main objectives for this analysis were to generate pooled estimates of the main effects of SQ-LNS on severe wasting and severe stunting and identify study-level modifiers of the effect of SQ-LNS on these outcomes.

## Methods

The protocol for the IPD meta-analysis was registered as PROSPERO CRD42019146592 (https://www.crd.york.ac.uk/prospero) (27). The detailed protocol was posted to Open Science Framework (https://osf.io/ymsfu) prior to analysis and updated after consultations with co-investigators before finalizing the analysis plan (28), and the results are reported according to PRISMA-IPD guidelines (29). The analyses were approved by the institutional review board of the University of California Davis (1463609–1). All individual trial protocols were approved by the relevant institutional ethics committees. The methods were presented in detail previously (23), so are summarized here.

### Inclusion and exclusion criteria for this IPD meta-analysis

We included randomized controlled trials of SQ-LNS provided to children age 6-24 months that met the following study-level inclusion criteria: 1) the trial was conducted in a low- or middle-income country (30); 2) SQ-LNS (< ∼125 kcal/d) was provided to the intervention group for at least 3 months between 6 and 24 months of age; 3) at least one trial group did not receive SQ-LNS or other type of child supplementation; 4) the trial reported at least one outcome of interest; and 5) the trial used an individual or cluster randomized design in which the same participants were measured at baseline (prior to child supplementation) and again after completion of the intervention (longitudinal follow-up), or different participants were measured at baseline and post-intervention (repeated cross-sectional data collection). Trials were excluded if: 1) only children with severe or moderate malnutrition were eligible to participate (i.e., SQ-LNS was used for treatment, not prevention of malnutrition); 2) the trial was conducted in a hospitalized population or among children with a pre-existing disease; or 3) SQ-LNS provision was combined with additional supplemental food or nutrients for the child within a single arm (e.g., SQ-LNS + food rations vs. control), and there was no appropriate comparison group (e.g., food rations alone) that would allow separation of the SQ-LNS effect from effects of the other food or nutrients provided.

Trials in which there were multiple relevant SQ-LNS interventions (e.g., varying dosages or formulations of SQ-LNS in different arms), combined provision of child SQ-LNS with provision of maternal LNS, or included other non-nutritional interventions (i.e., water, sanitation and hygiene (WASH)) were eligible for inclusion. In such trials, all arms that provided child SQ-LNS were combined into one group, and all non-LNS arms (i.e., no LNS for mother or child) were combined into a single comparator group for each trial (herein labeled “control”), excluding intervention arms that received non-LNS child supplementation (e.g., MNP, fortified-blended food). We also conducted a sensitivity analysis restricting the comparison to specified contrasts of intervention arms within multiple intervention trials (see below).

At the individual participant level, we included children if their age at baseline allowed them to receive at least 3 mo of intervention (supplementation or control group components) between 6 and 24 mo of age. We considered 3 mo to be the minimum duration for an impact on linear growth.

### Search methods and identification of studies

We identified studies cited in a previous systematic review and meta-analysis of child LNSs (26) and through keyword and controlled vocabulary searches of 25 databases, as described in Dewey et al.(23).

### Data collection

We invited all principal investigators of eligible trials to participate in the IPD meta-analysis. We provided a data dictionary listing definitions of variables requested for pooled analysis. Those variables were provided to the IPD analyst (CDA) in de-identified individual participant datasets.

### IPD integrity

We conducted a complete-case intention to treat analysis (31). We calculated length-for-age z-score (LAZ), weight-for-length z-score (WLZ), and mid-upper arm circumference z-score (MUACZ) using the 2006 WHO child growth standards and checked the values for acceptable standard deviations and to be within published WHO acceptable ranges (32). Biologically implausible values were flagged, as recommended by WHO, in the following way: LAZ <-6 or >6; WLZ<-5 or >5; MUACZ <-5 or >5. These were inspected for errors and either winsorized (33) if anthropometric values were biologically plausible or removed from analysis if values were clearly impossible. Such cleaning was necessary for less than 0.5% of participants, with a consistently low rate of implausibility across outcomes and studies. We also checked summary statistics, such as means and standard deviations, in our dataset against published values for each trial.

### Assessment of risk of bias in each study and quality of evidence across studies

Two independent reviewers (KRW and CDA) assessed risk of bias in each trial using the criteria outlined in the Cochrane Handbook for Systematic Reviews of Interventions (34). The same reviewers also assessed the quality of evidence for anthropometric outcomes across all trials based on the five GRADE criteria: risk of bias, inconsistency of effect, imprecision, indirectness, and publication bias (35).

### Specification of outcomes and effect measures

The statistical analysis plan pre-specified severe acute malnutrition (SAM) as an outcome (28) but we did not report it in our previous publication (23) because it is a rare event and thus poses difficulties for effect modification analysis. We updated the analysis plan to include the 4 outcomes reported herein, and added a sensitivity analysis to check the robustness of main effect estimates that exclude trials with zero events in at least one comparison group. The 4 outcomes are: severe wasting (WLZ < −3 SD), severe stunting (LAZ < −3 SD), SAM (WLZ < −3 SD or MUAC < 115 mm) and very low MUAC (MUACZ < −3 SD or MUAC < 115 mm). The main focus of this analysis is on severe wasting and severe stunting because sample sizes available for SAM and very low MUAC were considerably smaller. In addition, we were unable to include bilateral pitting edema as one of the criteria for SAM because this information was not collected in 5 of the trials, and in the other 9 trials the definition of edema varied (of those trials, 4 reported zero cases, 4 reported <20 cases, and 1 reported 38 cases of edema). For all 4 outcomes, the principal measure of effect was the prevalence ratio (PR) at endline.

The treatment of interest was provision of children with SQ-LNS (< ∼125 kcal/d, with or without co-interventions), compared to no intervention or an intervention without any type of LNS or other child supplement. Other types of interventions were delivered with or without LNS, such as WASH interventions and child morbidity monitoring and treatment. In several trials, child LNS was delivered to children whose mothers received maternal LNS during pregnancy and lactation. As described previously (23), we decided that if the main effects did not differ between the child-LNS-only analysis and the all-trials analysis (including maternal plus child LNS arms) by more than 0.05 for prevalence ratios, the results of the all-trials analyses would be presented as the principal findings, in order to maximize sample size. Three additional pre-specified sensitivity analyses were also conducted, as described below.

### Synthesis methods and exploration of variation in effects

We examined full sample main effects of the intervention for all 4 outcomes and evaluated whether certain characteristics modified the effects of SQ-LNS on severe wasting or severe stunting. The effect modification analyses focused on study-level characteristics. To be consistent with our previous publications we also examined potential individual-level effect modifiers, but those analyses were considered exploratory because some subgroups have zero “events” for rare outcomes such as severe wasting, which reduces the number of comparisons available for such outcomes. We used a two-stage approach for all analyses, which is preferred when incorporating cluster-randomized trials (36). In the first stage, we generated intervention effect estimates within each individual study according to its study design. For longitudinal study designs we controlled for initial child anthropometric status (at baseline or at the start of supplementation if enrollment occurred during pregnancy) when estimating the intervention effect on each outcome, to gain efficiency. To deal with outcome dependence in cluster-randomized trials, we used robust standard errors with randomization clusters as the independent unit. In the second stage, we pooled the first stage estimates using inverse-variance weighted fixed effects. We also conducted sensitivity analyses in which we pooled estimates using inverse variance weighted random effects (37, 38).

To evaluate main effects, we first estimated the intervention effect for each study. We then pooled the first stage estimates to generate a pooled point estimate, 95% confidence interval, and corresponding p-value. For effect modification analyses, we examined the dichotomous variables shown in **Supplemental Box 1**, as described previously (23). For study-level characteristics, we used random effects meta-regression to test the association between each effect modifier and the intervention. For individual-level characteristics, we generated pooled intervention effect estimates within each category to determine how the intervention effect in one subgroup differed from the intervention effect in the specified reference subgroup.

Heterogeneity of effect estimates was assessed using I^2^ and Tau^2^ statistics, within strata when relevant (39). We used a p-value of < 0.05 for main effects and a p-diff (from the random effects meta-regressions with study-level characteristics) or p-for-interaction (for individual-level characteristics) of < 0.10 for effect modification. Given that the growth outcomes are highly correlated and the effect modification analyses are inherently exploratory, we did not adjust for multiple hypothesis testing because doing so may be unnecessary and counterproductive (40).

For descriptive purposes, we calculated the number needed to avert a single case of severe wasting (“number needed to treat”, NNT) following the standard approach (41). The equation requires an assumed population prevalence of severe wasting among the untreated, and then the prevalence of severe wasting among the treated is estimated as the prevalence among the untreated multiplied by the prevalence ratio reduction for severe wasting. These two prevalences are then subtracted from one another and inverted. We repeated this calculation for various population prevalences that reflected the range of prevalence of severe wasting in the control groups in the actual trials, to understand how NNT would vary by context.

### Additional sensitivity analyses

As described previously (23), we conducted several pre-specified sensitivity analyses:

1. Separate comparisons within multi-component intervention trials, such that the SQ-LNS vs. no SQ-LNS comparisons were conducted separately between pairs of arms with the same non-nutrition components (e.g. SQ-LNS+WASH vs. WASH; SQ-LNS vs. Control). IYCF behavior change communication was not considered an additional component.
2. Exclusion of passive control arms, i.e., when control group participants received no intervention and had no contact with project staff between baseline and endline.
3. Exclusion of intervention arms with SQ-LNS formulations that did not include both milk and peanut.

In addition, we conducted a fourth sensitivity analysis, the “rare events” analysis, in which we included comparisons with zero-event-rates within an intervention arm for main effect analysis, to maximize the number of trials included. The primary analytic approach did not produce effect estimates for trials with zero events in at least one arm (3 trials for severe wasting, 1 trial for SAM, 2 trials for very low MUAC), so an estimate was generated for those trials by adapting the approach described in the Cochrane Handbook for Systematic Reviews of Interventions (34) where an event count of 0.5 is substituted for the 0 event count value observed in the trial.

## Results

### Literature search and trial characteristics

We identified 15 trials that met our inclusion criteria, 14 of which provided individual participant data and are included in this analysis **(Table 1, Supplemental Figure 1, Supplemental Table 1)** (42–56). Investigators for one trial were unable to participate (57); binary outcomes were not reported in that trial, so we were unable to insert an estimate from that trial into our analysis. One trial was designed *a priori* to present results separately for HIV-exposed and HIV-unexposed children, so we present it as two separate comparisons (55, 56). Similarly, the two PROMIS trials in Burkina Faso and Mali each included an independent longitudinal cohort and repeated (at baseline and endline) cross-sectional samples, so the longitudinal and cross-sectional results are presented as separate comparisons (46, 54). Thus, the 14 trials yielded 17 separate comparisons.

**Table 1.**
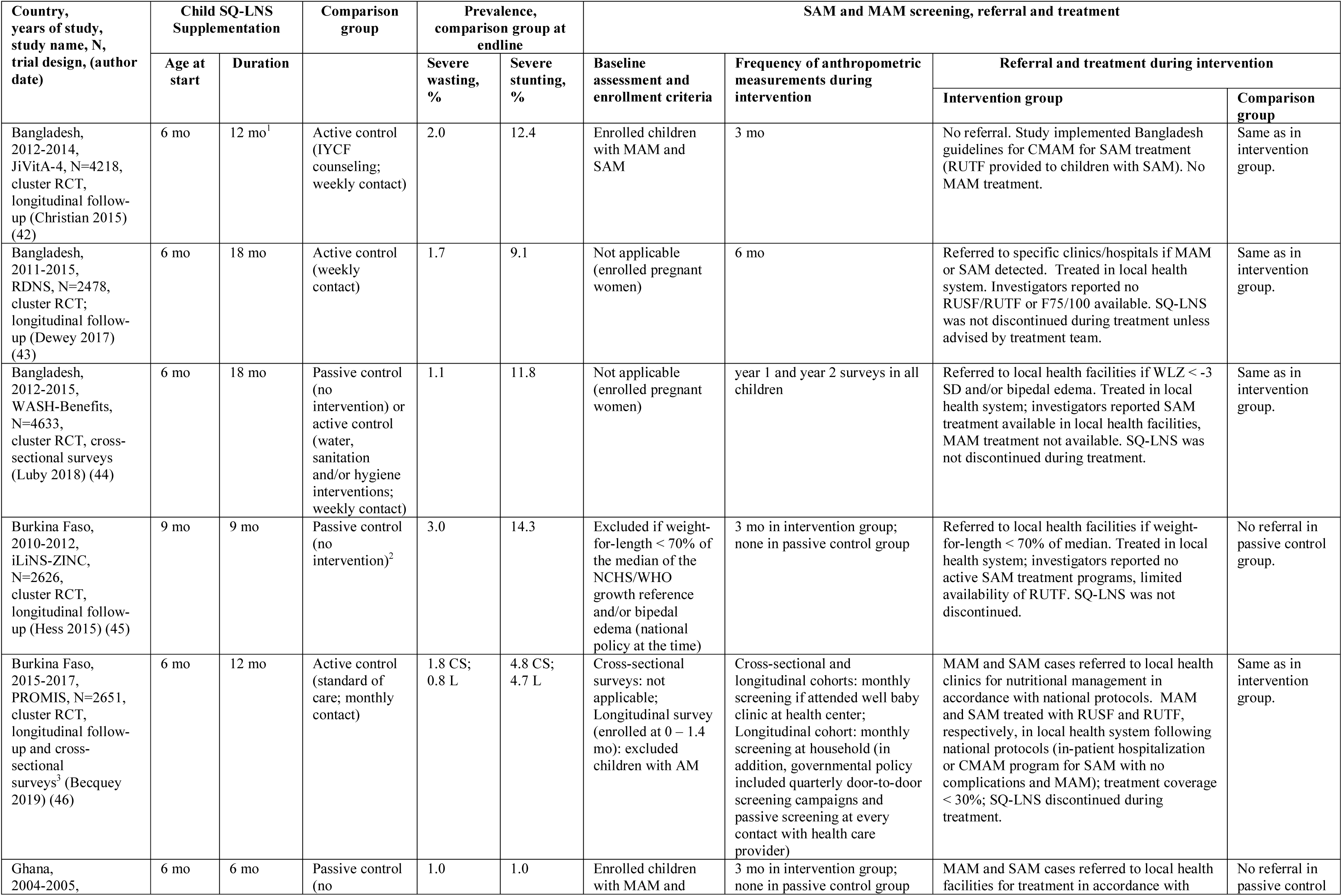
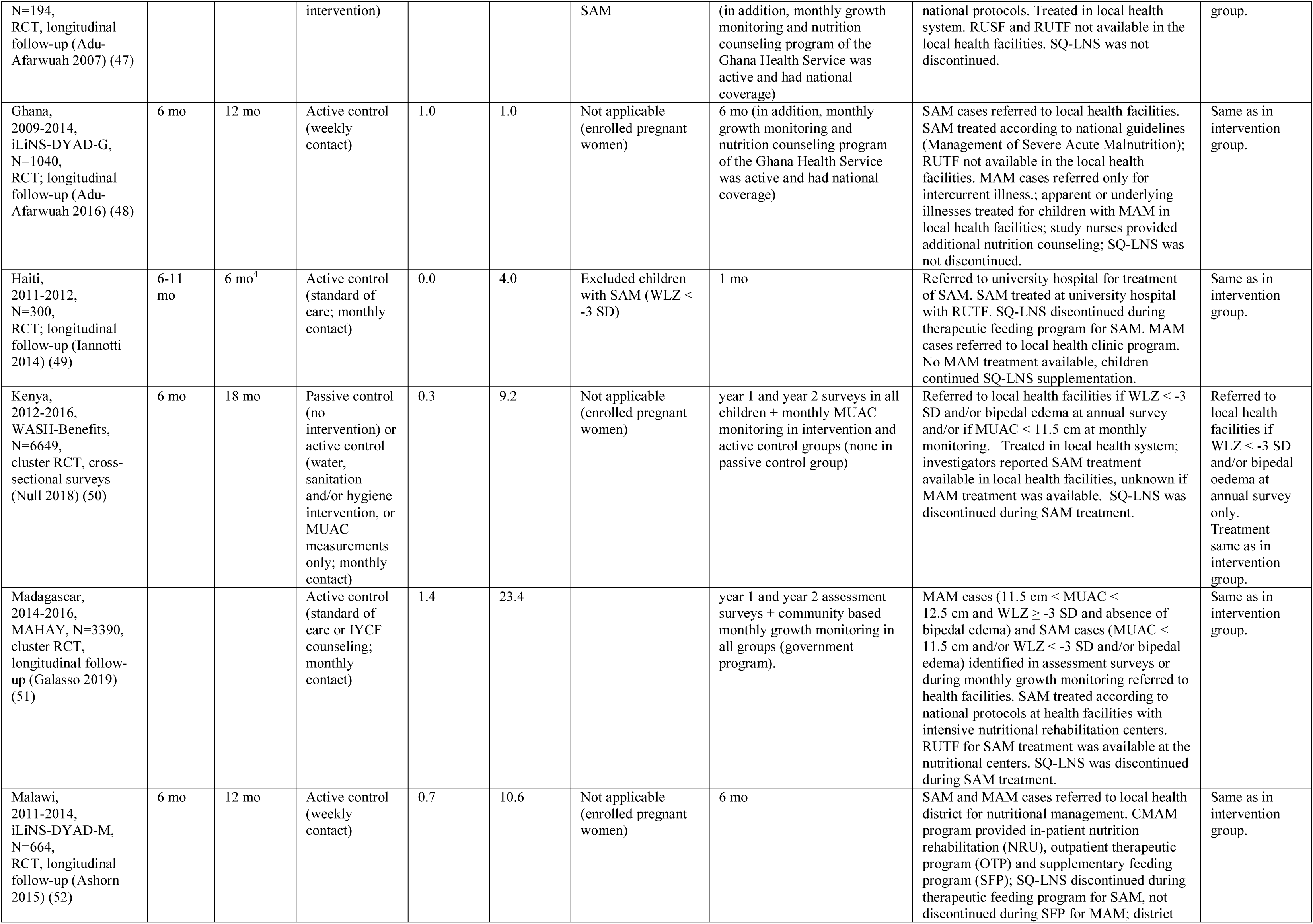
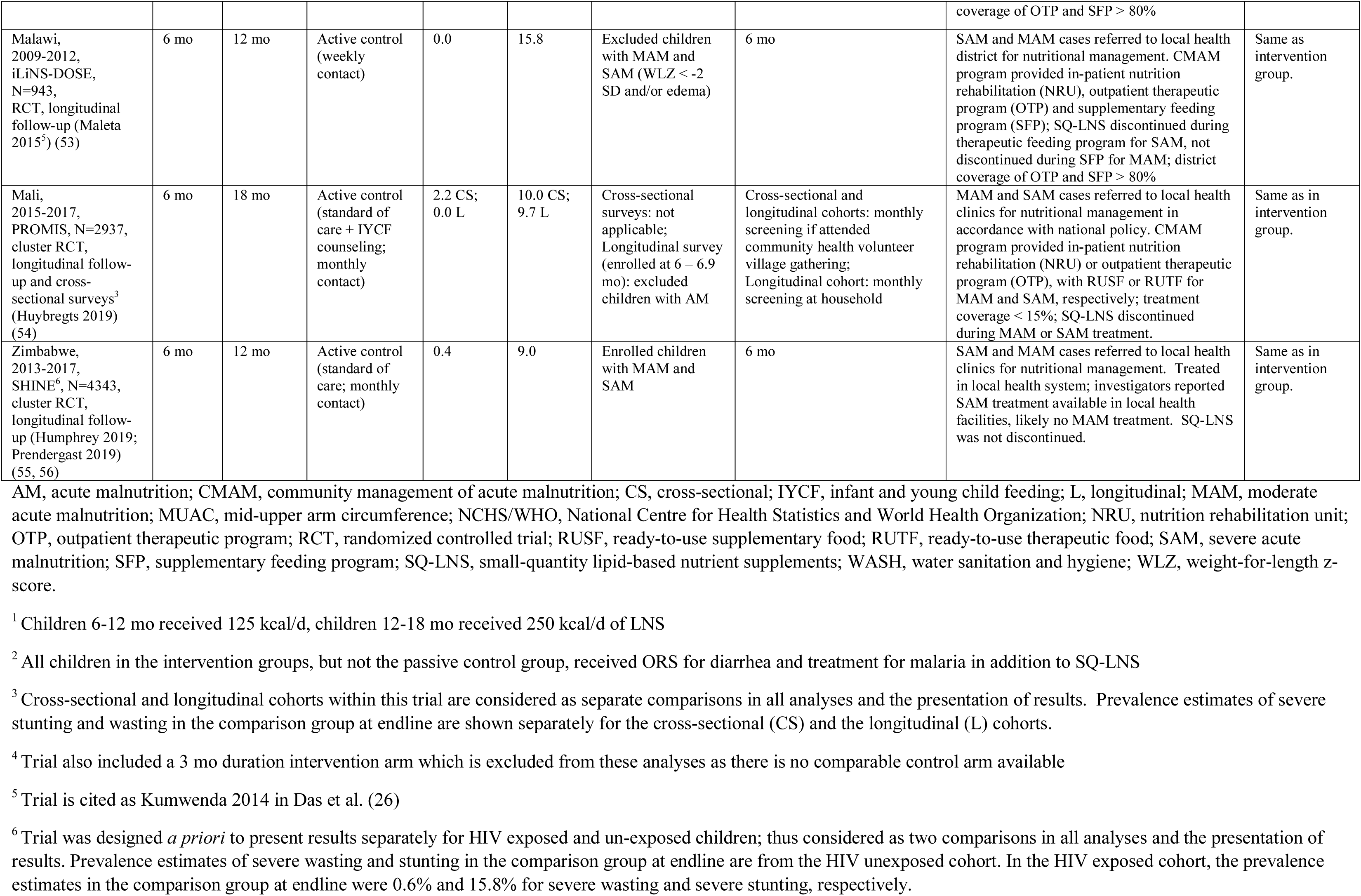
Characteristics of trials included in the individual participant data analysis.

The 14 trials in these analyses were conducted in Sub-Saharan Africa (10 trials in 7 countries), Bangladesh (3 trials), and Haiti (1 trial), and included a total of 37,066 infants and young children with anthropometric data. The majority of trials began child supplementation with SQ-LNS at 6 months of age and the intended duration ranged from 6 to 18 months of supplementation. The SQ-LNS for children generally provided approximately 120 kcal/d and ∼1 RDA of 19-22 micronutrients (23); in one trial the ration was ∼120 kcal/d between 6 and 12 months of age and ∼250 kcal/d between 12 and 24 months of age (42). Six trials were conducted within existing community-based or clinic-based programs (43, 46, 49, 51, 54–56); in the other trials, all activities were conducted by research teams. All trials provided social and behavior change communication (SBCC) on IYCF to reinforce the normal IYCF messages already promoted in that setting or to go beyond the usual IYCF messaging (23), in addition to information on how to use SQ-LNS for the target child. Three trials included arms with WASH interventions (44, 50, 55, 56). Most trials provided comparisons that included an active control arm (i.e., similar contact frequency as for intervention arms) but 2 were limited to comparisons with a passive control arm (45, 47).

There was variability across trials with regard to screening, referral and treatment for SAM and moderate acute malnutrition (MAM) among participants (Table 1). In some trials, acute malnutrition at baseline was an exclusion criterion (45, 46, 49, 53, 54), though the definition used for acute malnutrition varied, while other trials did not exclude children with SAM or MAM (42, 46, 47, 51, 54–56), or enrollment occurred during pregnancy and thus no such exclusion criteria were applicable (43, 44, 48, 50–52). Once enrolled, most trials included anthropometric assessments of participants in both intervention and control arms on a regular basis (monthly (46, 49, 50, 54); every 3 mo (42); or every 6 mo (43, 48, 52, 53, 55, 56)). In 2 trials, measurements were conducted only during yearly surveys, in both intervention and control groups (44, 51), and in 3 trials measurements occurred monthly (50) or every 3 mo (45, 47) in the intervention group (and active control group in WASH-B Kenya) but not in the (passive) control group. For children identified with SAM or MAM, 1 trial provided treatment for SAM (but not MAM) directly to participants (42), and all other trials referred malnourished children to local health facilities for treatment, though the criteria for referral varied. In some sites, treatment for MAM via local programs was available (though coverage may have been low) (46, 52–54), but in most sites MAM treatment was unavailable or unlikely. SAM treatment, however, was reportedly offered in most sites, although referral follow-through and/or availability of ready-to-use-therapeutic food may have been limited.

The prevalences of severe wasting and severe stunting in the control groups at endline are shown in Table 1. Prevalence of severe wasting ranged from 0% in Haiti and Malawi (iLiNS-DOSE trial) to 3% in Burkina Faso (iLiNS-ZINC trial). Prevalence of severe stunting ranged from 1% in Ghana to 23% in Madagascar. Additional descriptive information on study-level characteristics is presented in **Supplemental Table 2**. At the study level, 6 sites had a high burden of wasting (≥ 10% in the control group at endline: Mali, both sites in Burkina Faso, and all 3 sites in Bangladesh) and 8 of the 14 study sites had a high burden of stunting (≥ 35% in the control group at 18 mo). Country-level malaria prevalence ranged from <1% in Bangladesh and Haiti to 59% in Burkina Faso. Study-specific prevalence of improved water quality ranged from 27% to 100%, and prevalence of improved sanitation ranged from 0% to 97%. Frequency of contact during the study was weekly in 7 trials and monthly in 7 trials. Average estimated reported compliance with SQ-LNS consumption was categorized as high (≥80%) in 7 trials and ranged between 37% and 77% in the other trials. Individual-level characteristics were reported previously (23).

As reported previously (23), we considered the trials to have a low risk of bias for most of the criteria (**Supplemental Figure 2**). For blinding of participants, all trials were judged to have high risk of bias, as blinding was not possible given the nature of the intervention.

### Main effects of SQ-LNS

SQ-LNS reduced the prevalence of adverse growth outcomes by 31% for severe wasting, 17% for severe stunting, 24% for SAM and 27% for very low MUAC (**Table 2**, **Figures 1** and **2**, **Supplemental Figures 3 and 4)**. Results from the child-LNS-only and all-trials analyses were similar: for all 4 outcomes, the PRs for intervention vs control groups were identical or almost identical when the maternal LNS trials/arms were included (**Figure 3**, **Supplemental Figures 5 and 6**). Therefore, results from the all-trials analyses, inclusive of maternal + child LNS trials/arms, are presented as the principal findings. For all outcomes, fixed-effects and random-effects models generated identical or very similar estimates. We rated the quality of the evidence for all outcomes as high based on GRADE criteria: at least 10 randomized controlled trials were available for all outcomes, risk of bias was low, heterogeneity was generally low to moderate (Table 2), precision was rated as high because all but 2 trials had sample sizes > 600, all trials were directly aimed at evaluating SQ-LNS, and funnel plots revealed no indication of publication bias (35).

**Figure 1:**
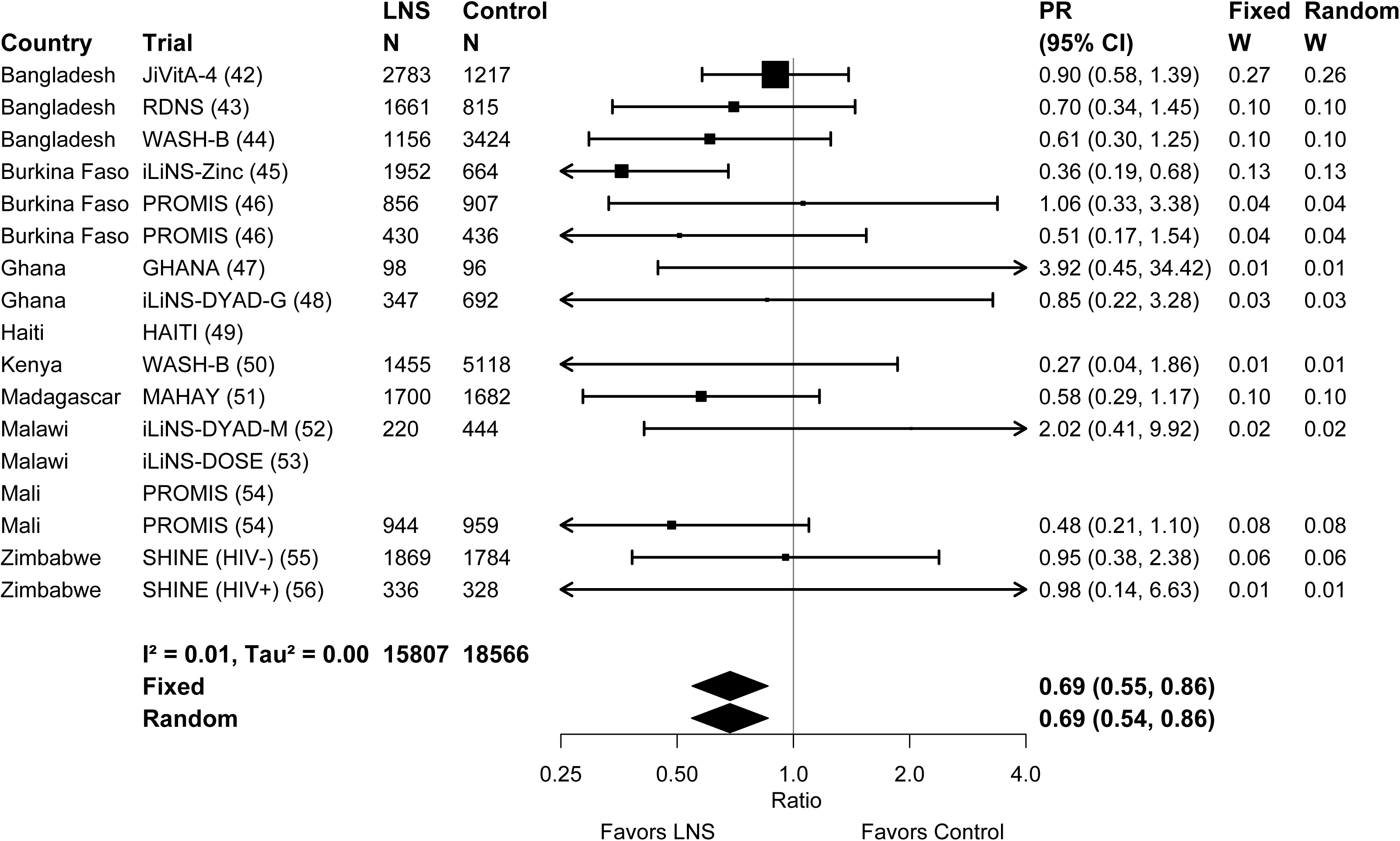
Forest plot of effect of SQ-LNS on severe wasting prevalence. LNS, lipid-based nutrient supplements; PR, prevalence ratio. Individual study estimates were generated from log- binomial regression controlling for baseline measure when available and with clustered observations using robust standard errors for cluster-randomized trials. Pooled estimates were generated using inverse-variance weighting with both fixed and random effects.

**Figure 2:**
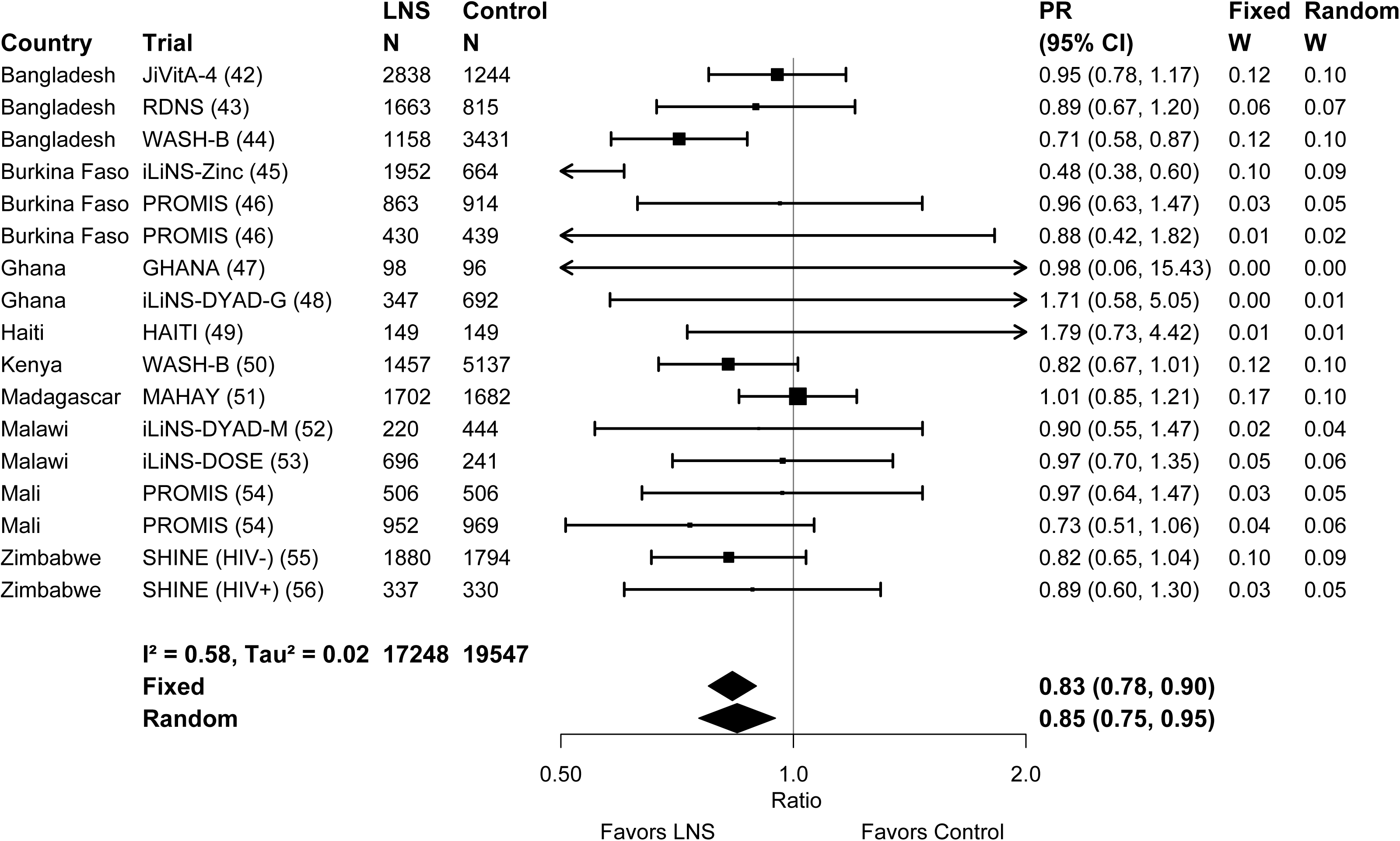
Forest plot of effect of SQ-LNS on severe stunting prevalence. LNS, lipid-based nutrient supplements; PR, prevalence ratio. Individual study estimates were generated from log- binomial regression controlling for baseline measure when available and with clustered observations using robust standard errors for cluster-randomized trials. Pooled estimates were generated using inverse-variance weighting with both fixed and random effects.

**Figure 3:**
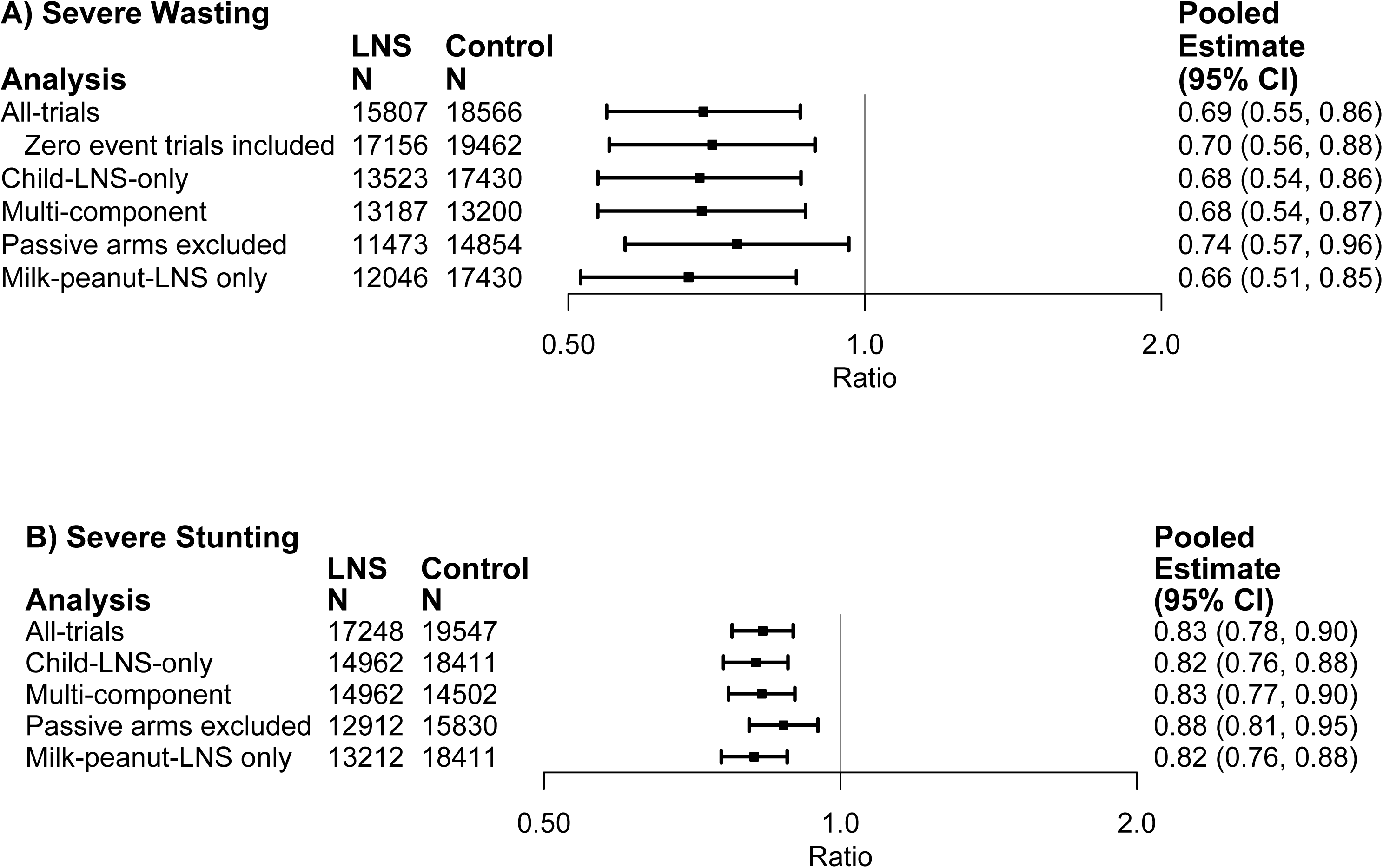
Sensitivity analyses of main effects of SQ-LNS on prevalence ratios for severe stunting (A) and severe wasting (B). All-trial analysis includes all trials; Child-LNS-only excludes trial arms that provided both maternal and child LNS; Multi-component analysis separates comparisons within trials that included multi-component interventions, so that the SQ-LNS vs. no SQ-LNS comparisons were conducted separately between pairs of arms that included the same non-nutrition components (e.g. SQ-LNS+WASH vs. WASH; SQ-LNS vs. Control); Passive arms excluded analysis excludes passive control arms; Milk-peanut LNS only analysis excludes arms with SQ-LNS formulations that were not milk and peanut based; “Zero events trials included” uses estimates generated for certain trials in which 0.5 is substituted for the 0 value in analysis for severe wasting.

**Figure 4:**
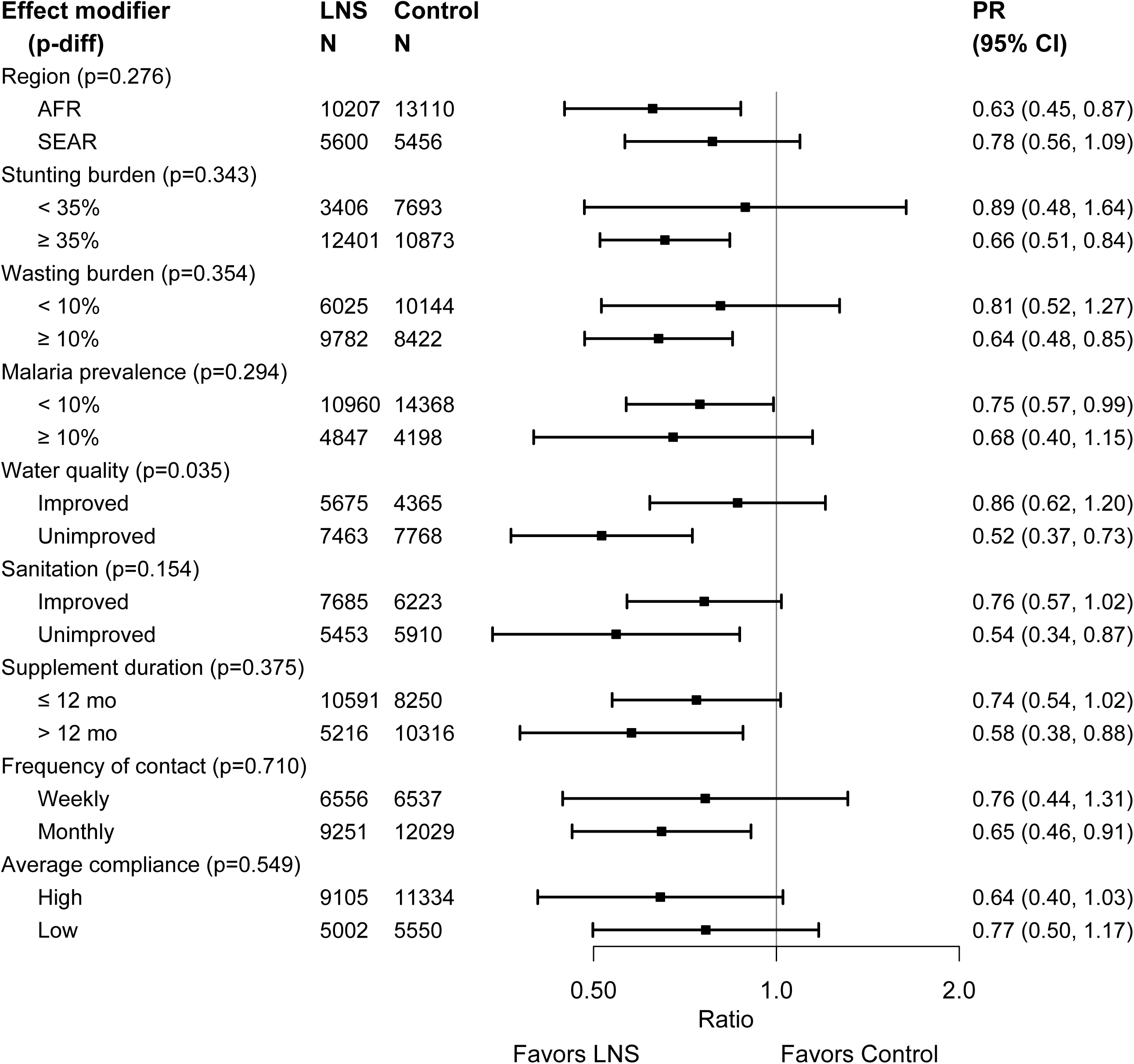
Pooled effect of SQ-LNS on severe wasting stratified by study-level characteristics. AFR, African Region; LNS, lipid-based nutrient supplements; P-diff, p-value for the difference in effects of SQ-LNS between the two levels of the effect modifier; PR, prevalence ratio; SEAR, South-East Asia Region. P-value for the difference was estimated using random effects meta- regression with the indicated effect modifier as the predictor of intervention effect size; stratified pooled estimates are presented for each strata.

**Figure 5:**
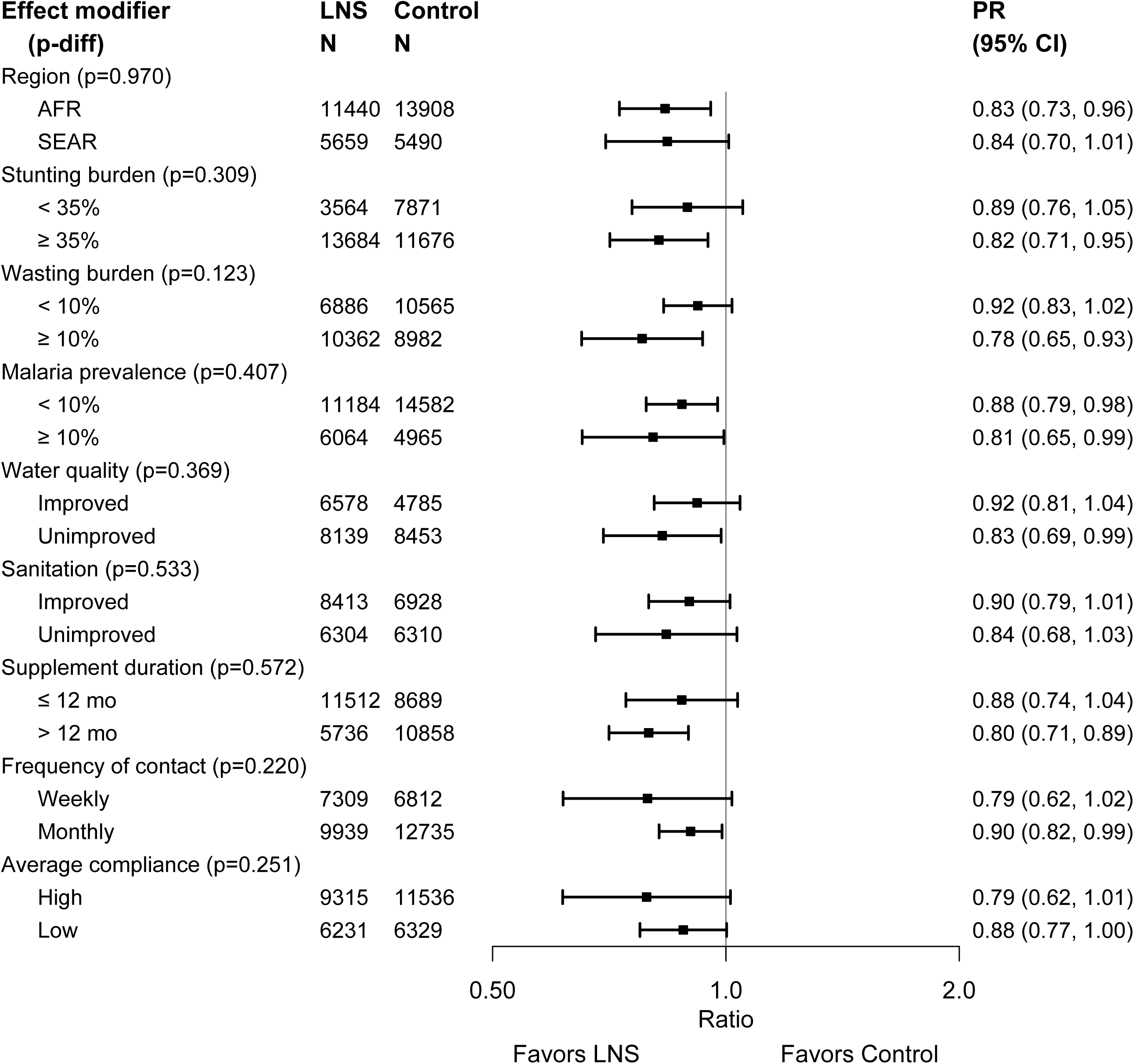
Pooled effect of SQ-LNS on severe stunting stratified by study-level characteristics. AFR, African Region; LNS, lipid-based nutrient supplements; P-diff, p-value for the difference in effects of SQ-LNS between the two levels of the effect modifier; PR, prevalence ratio; SEAR, South-East Asia Region. P-value for the difference was estimated using random effects meta- regression with the indicated effect modifier as the predictor of intervention effect size; stratified pooled estimates are presented for each strata.

**Table 2.** Main effects of SQ-LNS on severe wasting, severe stunting, severe acute malnutrition and very low MUAC.

| **Outcomes** | **Number of participants (comparisons)** | **Prevalence ratio SQ-LNS vs Control (95% CI)** | **P value<sup>1</sup>** | **Heterogeneity I<sup>2</sup> (p-for-heterogeneity)<sup>2</sup>** | **Grade** |
| --- | --- | --- | --- | --- | --- |
| Severe Wasting (WLZ < -3 SD) | 34373 (14) | 0.69 (0.55, 0.86) | 0.001 | 0.01 (0.856) | High |
| Severe Stunting (LAZ < -3 SD) | 36795 (17) | 0.83 (0.78, 0.90) | <0.001 | 0.58 (<0.001) | High |
| Severe Acute Malnutrition (WLZ < -3 SD or MUAC < 115 mm) | 30436 (13) | 0.76 (0.62, 0.93) | 0.008 | 0.00 (0.892) | High |
| Very low MUAC (MUACZ < -3 SD or < 115 mm) | 30069 (12) | 0.73 (0.56, 0.94) | 0.015 | 0.24 (0.609) | High |

LAZ, length-for-age z-score; LNS, lipid-based nutrient supplements; MUAC, mid-upper arm circumference; MUACZ, mid-upper arm circumference z-score; SQ-LNS, small-quantity lipid-based nutrient supplements; WLZ, weight-for-length z-score.
<sup>1</sup>The P Value column corresponds to the pooled main effect two-sided superiority testing of the intervention effect estimate and 95% confidence interval presented in the preceding column.
<sup>2</sup>I<sup>2</sup> describes the percentage of variability in effect estimates that may be due to heterogeneity rather than chance. Roughly, 0.3 to 0.6 may be considered moderate heterogeneity. P-value from chi-squared test for heterogeneity. P < 0.05 indicates statistically significant evidence of heterogeneity of intervention effects beyond chance.

Results were similar in most of the sensitivity analyses (Figure 3, Supplemental Figures 5 and 6), including the “rare events” sensitivity analysis (forest plot shown for severe wasting in **Supplemental Figure 7**). However, there was some attenuation of the effects when passive control arms were excluded; e.g., the PR for severe wasting was 0.74 (0.57, 0.96) and the PR for severe stunting was 0.88 (0.81, 0.95).

The number of children who would need to be provided with SQ-LNS to prevent 1 case of severe wasting (NNT) varies depending on the estimated prevalence of severe wasting in the study area, as shown in **Supplemental Table 3**. The average prevalence of severe wasting in the control groups at endline was ∼1%. At this prevalence, the NNT is 323 assuming a relative reduction of 31% or 385 assuming a relative reduction of 26%. In sites with a higher prevalence, e.g., 3%, the NNT is 108 or 128, assuming relative reductions of 31% or 26%, respectively. If the NNT estimate is based on longitudinal data such as incidence of severe wasting during a 12- mo period, which could be as high as 5-24% (7, 46, 54), the NNT ranges from 13 to 77.

### Effect modification

Forest plots for severe wasting and severe stunting stratified by study-level effect modifiers are presented in **Supplemental Figures 8A-I and 9A-I,** and results are summarized in **Figures 4 and 5**. For severe wasting, there was a significantly greater effect of SQ-LNS in sites with unimproved water quality (PR 0.52 (0.37, 0.73)) than in sites with better water quality (PR 0.86 (0.62, 1.20), p-diff = 0.035). For severe stunting, none of the tests for effect modification was statistically significant. For both outcomes, in many cases the differences in prevalence ratios between strata were sizable (e.g., > 0.10) even though the p-diff for interaction was not significant, presumably due to limited statistical power for these types of analyses. For example, the PR for severe stunting was 0.78 (0.65, 0.93) in sites with a wasting burden ≥ 10%, compared to 0.92 (0.83, 1.02) in sites with a lower wasting burden. For severe wasting, notable differences between strata (apart from the water quality interaction noted above) were evident for region (PR 0.63 (0.45, 0.87) for African sites; 0.78 (0.56, 1.09) for Bangladesh sites), stunting burden (PR 0.66 (0.51, 0.84) in high stunting burden sites vs. 0.89 (0.48, 1.64) in lower stunting burden sites), wasting burden (PR 0.64 (0.48, 0.85) in high wasting burden sites vs. 0.81 (0.52, 1.27) in lower wasting burden sites), and sanitation (PR 0.54 (0.34, 0.87) in sites with unimproved sanitation vs. 0.76 (0.57, 1.02) in site with better sanitation).

In the exploratory analysis of the potential individual-level modifiers, only a few characteristics significantly modified the effect of SQ-LNS on severe wasting or severe stunting (**Supplemental Table 4**). As expected, for some characteristics, the number of comparisons available for analysis of effect modification was greatly reduced from the numbers available for analysis of main effects (particularly for severe wasting), so statistical power may be limited. Of the 28 interactions examined (14 individual-level characteristics x 2 outcomes), only 2 (7%) met the criterion of p-for-interaction < 0.10, which is what could be expected due solely to chance. However, in one case the p-for-interaction was < 0.0001, which is less likely to be due to chance: there was a greater effect of SQ-LNS on severe stunting among later-born children (PR 0.77 (0.71, 0.84)) than among first-born children (PR 0.94 (0.84, 1.06)).

## Discussion

In this large IPD analysis (n ∼37,000), the relative reductions in severe adverse growth outcomes following provision of SQ-LNS to infants and young children 6 to 24 months of age were 31% for severe wasting and 17% for severe stunting. Results were similar regardless of inclusion/exclusion of arms with maternal plus child SQ-LNS, or arms with non-standard SQ-LNS formulations, as well as when analyses of multi-component intervention trials were structured to more specifically isolate the effects of SQ-LNS. Effects were attenuated, though still significant, when comparisons using passive control arms were excluded, with relative reductions of 26% for severe wasting and 12% for severe stunting. Effects of SQ-LNS appeared to be greater in sites with greater burdens of stunting or wasting, or with poorer water quality or sanitation, although the only statistically significant study-level effect modifier was water quality: the relative reduction in severe wasting was 48% in sites with unimproved water quality, compared to 14% in sites with better water quality.

Our estimate of the effect of SQ-LNS on severe stunting (17% relative reduction) is similar to the estimated 15% relative reduction reported by Das et al. (26), based on 6,151 children in 5 studies that included both SQ-LNS and medium-quantity LNS. In their meta-analysis, there was no effect on severe wasting but there were only 3 studies and fewer than 2,500 children. The relatively large reduction in the prevalence of severe wasting in our IPD analysis, restricted to trials that used SQ-LNS, is thus a novel finding of considerable global health significance and relevant to current initiatives aimed at preventing and treating wasting (11–13). Attenuation of the effect when comparisons using passive control arms were excluded (from a 31% to a 26% relative reduction in severe wasting) is consistent with the results of a previous meta-analysis of effects of LNS (mostly SQ-LNS) on all-cause mortality from 6 to 24 months of age (58). Part of the impact of an SQ-LNS intervention on severe wasting or mortality, when a passive control arm is the comparator, could be due to more frequent contact with a health worker or data collector, which could lead to greater care for the child as well as detection and treatment of acute malnutrition. Nonetheless, the protective effect of SQ-LNS on severe wasting prevalence is substantial even if this potential phenomenon is taken into account. In areas with a relatively high burden of severe wasting, the number of children who would need to be provided with SQ-LNS to prevent 1 case of severe wasting, estimated based on a cross-sectional prevalence of 3%, would be ∼108 assuming a relative reduction of 31% and ∼128 assuming a relative reduction of 26%. If estimated based on a 15% incidence of severe wasting over a 12-mo period, the NNT would be ∼22 or 26 assuming a relative reduction of 31% or 26%, respectively.

The estimated reduction in severe wasting due to SQ-LNS in our pooled analyses captures only the impact on prevalence at endline, not on longitudinal prevalence or incidence. In Mali, the SQ-LNS intervention had no effect on the prevalence of SAM in the cross-sectional sample of children but reduced the longitudinal prevalence of SAM in the longitudinal cohort by 43% though some of this difference could be due to the season in which the cross-sectional sample was assessed (54). Another consideration is that children who died during the study period did not enter into our calculation of the estimated prevalence of severe wasting at endline. Mortality was lower in the SQ-LNS arms (58), and if severe wasting was associated with mortality then more severely wasted children could be “missing” from the control arm because they died and were excluded from analysis, which would lead to an underestimate of the effect of SQ-LNS on severe wasting.

The pooled estimates for relative reductions in severe wasting and severe stunting, across all 14 trials, may also be underestimates of potential effects of SQ-LNS in the highest-risk populations. Effect modification by study-level characteristics was generally not statistically significant, but statistical power for these analyses was constrained by the limited number of trials. As a result, there may be meaningful differences in effect estimates between categories of trials even if the p- diff for the association between the effect modifier and effect size was not significant. For example, the relative reduction in severe stunting due to SQ-LNS was 22% in sites with a wasting burden ≥ 10%, compared to 8% in sites with a lower wasting burden. For severe wasting, the relative reduction due to SQ-LNS was 36% in sites with a high wasting burden, 34% in sites with a high stunting burden, 46% in sites with unimproved sanitation and 48% in sites with unimproved water quality. These findings suggest that targeting preventive SQ-LNS to high-risk populations may be warranted, which is consistent with the IPD analysis results for developmental outcomes (24) and hemoglobin (25).

The effects on severe wasting reported herein need to be interpreted in the context of how the trials handled children with acute malnutrition at baseline or thereafter. Most of the trials did not exclude children with MAM or SAM from participating (e.g., several trials enrolled during pregnancy), so the results should be generalizable to the general population in those sites. However, most trials did include regular anthropometric assessments of children during the study period and provided treatment or referred children for treatment of acute malnutrition (mainly for SAM, as MAM treatment was less likely to be available locally). Thus, by the time of the endline assessment, such children may no longer have been severely wasted, which could have biased our effect estimates towards the null. For most trials, assessment, referral and treatment of children with acute malnutrition was the same in both the intervention and control arms, except for the 2 studies in which no active control arm was included (45, 47). The sensitivity analysis excluding passive control arms accounts for the potential bias introduced by those differences.

Strengths of these analyses include the large sample size, the substantial number of high-quality randomized controlled trials available, and the high participation rate among investigators invited to contribute data. The 14 study sites were diverse in terms of geographic location, stunting burden, malaria prevalence, water quality, sanitation and several aspects of study design, which provided heterogeneity for exploration of study-level potential effect modifiers. Six of the 14 trials in this IPD analysis were conducted within existing community-based or clinic-based programs (43, 46, 49, 51, 54–56), so the findings include studies carried out in a real-world context. There are some limitations, however. Bangladesh was the only country represented in the Southeast Asia Region, and Haiti was the only country represented in Latin America and the Caribbean. Caution is needed when interpreting the effect modification results because statistical power was limited and many of the study-level characteristics are inter-related (e.g., sites with unimproved water quality also tended to have unimproved sanitation). Thus, attribution of differences in impact of SQ-LNS to a particular study-level characteristic may not be warranted.

These results add to the body of evidence demonstrating benefits of preventive SQ-LNS for infants and young children across multiple outcomes, including child growth (23), iron deficiency and anemia (25), child development (24) and child mortality (58). The effects on severe wasting and severe stunting demonstrated herein strengthen our previous recommendation that policymakers and program planners should consider including SQ-LNS in the mix of interventions to prevent adverse growth outcomes (23). They also provide more evidence for the potential mechanisms by which SQ-LNS reduces child mortality, i.e., via reductions in severe wasting and severe stunting. SQ-LNS is not a stand-alone intervention and should always be accompanied by messaging to reinforce recommended IYCF practices. When included in existing platforms for promoting healthy growth and development, such as community health worker programs, evidence is emerging to suggest that SQ-LNS may be a very cost-effective intervention in terms of costs per disability-adjusted life year (59). The effects on severe wasting are highly relevant to the goals of the United Nation’s Global Action Plan on Child Wasting (12), especially considering the paucity of evidence on effective strategies to prevent severe wasting. An important next step is additional cost-effectiveness analyses of incorporating SQ-LNS within integrated programs to prevent and treat wasting, taking into account the potential for reducing the number of cases of both moderate and severe wasting that would need treatment with supplemental or therapeutic foods, as well as reduced numbers of children requiring hospitalization.

## Supporting information

Supplemental Figures Table of Contents

Supplemental Box 1

PRISMA checklist

## Data Availability

Data described in the manuscript, code book and analytic code will not be made available because they are compiled from 14 different trials, and access is under the control of the investigators of each of those trials.

## Acknowledgments

We thank all of the co-investigators, collaborators, study teams, participants and local communities involved in the trials included in these analyses. These trials benefitted from the contributions of many partner organizations, including: icddr,b (JiVitA-4, Rang-Din Nutrition Study and WASH Benefits trial in Bangladesh); the World Food Program (JiVitA-4 trial in Bangladesh); the Health District of Dandé and the relevant local health-care authorities (iLiNS-ZINC trial in Burkina Faso); AfricSanté and Helen Keller International (PROMIS trials in Burkina Faso and Mali); Ministry of Public Health and Population (Haiti trial); Innovations for Poverty Action and the Kenya Medical Research Institute (WASH-Benefits trial in Kenya); Unité Programme National de Nutrition Communautaire, Government of Madagascar, and World Bank Health and Nutrition and Population Global Practice (MAHAY trial in Madagascar); the Ministry of Health and Child Care in Harare, Chirumanzu and Shurugwi districts, and Midlands Province (SHINE trial in Zimbabwe); the International Lipid-based Nutrient Supplements Project Steering Committee (iLiNS Project trials); and Nutriset (for development of SQ-LNS). We thank Emily Smith for advice on IPD analysis methods.

The authors’ responsibilities were as follows—KGD: drafted the manuscript with input from KRW, CDA, ELP, CPS and other coauthors; KRW, CDA, KGD, ELP and CPS: wrote the statistical analysis plan; BFA, PA, EB, LH and JHH: reviewed, contributed to, and approved the statistical analysis plan; KRW and CDA: compiled the data; CDA: conducted the data analysis; and all authors: read, contributed to, and approved the final manuscript; KGD is responsible for final content.

Supported by Bill & Melinda Gates Foundation grant OPP49817 (to KGD). KRW received a grant from Nutriset, SAS outside of the submitted work during the period of this IPD analysis project; BA received travel support (airfare and hotel) covered by the Bill & Melinda Gates Foundation to attend meetings in Seattle during the period of this IPD analysis project; PC was an employee of the Bill & Melinda Gates Foundation when this project was conceived until ber 2019. All other authors report no conflicts of interest.

## Abbreviations

AFR: African Region
IPD: individual participant data
IYCF: infant and young child feeding
LAZ: length-for-age z-score
LNS: lipid-based nutrient supplements
MAM: moderate acute malnutrition
MMN: multiple micronutrients
MNP: multiple micronutrient powder
MUAC: mid-upper arm circumference
MUACZ: mid-upper arm circumference z-score
NNT: number needed to treat
PR: prevalence ratio
RCT: randomized controlled trial
SAM: severe acute malnutrition
SBCC: social and behavior change communication
SEAR: South-East Asia Region
SQ-LNS: small-quantity lipid-based nutrient supplements
WASH: water sanitation and hygiene
WLZ: weight-for-length z-score.

## References

1. United Nations Children’s Fund (UNICEF), World Health Organization, International Bank for Reconstruction and Development/The World Bank. Levels and trends in child malnutrition: key findings of the 2021 edition of the joint child malnutrition estimates. Geneva: World Health Organization; 2021.

2. Stevens G, Finucane M, Paciorek G, Flaxman S, White R, Donner A, Ezzat M. Trends in mild, moderate, and severe stunting and underweight, and progress towards MDG 1 in 141 developing countries: a systematic analysis. Lancet 2012;380:824–34.

3. United Nations Children’s Fund (UNICEF). Guidance for estimating the number of children in need of treatment for wasting. New York: UNICEF Nutiriton; 2021.

4. Barba FM, Huybregts L, Leroy JL. Incidence Correction Factors for Moderate and Severe Acute Child Malnutrition From 2 Longitudinal Cohorts in Mali and Burkina Faso. Am J Epidemiol 2020;189:1623–7.

5. Wali N, K EA, Renzaho AMN. Wasting and Associated Factors among Children under 5 Years in Five South Asian Countries (2014-2018): Analysis of Demographic Health Surveys. Int J Environ Res Public Health 2021;18.

6. Headey DD, Ruel MT. Economic shocks predict increases in child wasting prevalence. Nat Commun 2022;13:2157.

7. Mertens A, Benjamin-Chung J, Colford Jr J, Hubbard A, van der Laan M, Coyle J, Sofrygin O, Cai W, Jilek W, Rosete S, et al. Child wasting and concurrent stunting in low- and middle-income countries. medRxiv 2020.06.09.20126979; 2021. doi: https://doi.org/10.1101/2020.06.09.20126979.

8. Olofin I, McDonald CM, Ezzati M, Flaxman S, Black RE, Fawzi WW, Caulfield LE, Danaei G, Nutrition Impact Model S. Associations of suboptimal growth with all-cause and cause-specific mortality in children under five years: a pooled analysis of ten prospective studies. PLoS One 2013;8:e64636.

9. Grey K, Gonzales GB, Abera M, Lelijveld N, Thompson D, Berhane M, Abdissa A, Girma T, Kerac M. Severe malnutrition or famine exposure in childhood and cardiometabolic non- communicable disease later in life: a systematic review. BMJ Glob Health 2021;6.

10. Waber DP, Bryce CP, Girard JM, Zichlin M, Fitzmaurice GM, Galler JR. Impaired IQ and academic skills in adults who experienced moderate to severe infantile malnutrition: a 40-year study. Nutr Neurosci 2014;17:58–64.

11. United Nations Children’s Fund (UNICEF). Severe wasting: an overlooked child survival emergency. UNICEF - Child Alert May 2022. Available at: https://www.unicef.org/child-alert/severe-wasting. Accessed on: 31 May 2022.

12. United Nations Children’s Fund, UN Food and Agriculture Organization, United Nations High Commissioner for Refugees, World Food Programme and the World Health Organization. Global action plan on child wasting: a framework for action to accelerate progress in preventing and managing child wasting and the achievement of the Sustainable Development Goals. New York, 2021. Available at: www.childwasting.org. Accessed on: 31 May 2022.

13. World Health Organization. WHO guideline development group meeting - scoping meeting for the WHO guideline on the prevention and treatment of wasting in infants and children. December 8–11, 2020. Available at: https://www.who.int/news-room/events/detail/2020/12/08/default-calendar/who-guideline-development-group-meeting-scoping-meeting-for-the-who-guideline-on-the-prevention-and-treatment-of-wasting-in-infants-and-children#:~:text=To%20achieve%20this%2C%20the%20SDGs,the%20world%20has%20remained%20unchanged. Accessed on: 31 May 2022.

14. Stewart CP, Iannotti L, Dewey KG, Michaelsen KF, Onyango AW. Contextualising complementary feeding in a broader framework for stunting prevention. Matern Child Nutr 2013;9 Suppl 2:27–45.

15. Prendergast AJ, Humphrey JH. The stunting syndrome in developing countries. Paediatr Int Child Health 2014;34:250–65.

16. Millward DJ. Nutrition, infection and stunting: the roles of deficiencies of individual nutrients and foods, and of inflammation, as determinants of reduced linear growth of children. Nutr Res Rev 2017;30:50–72.

17. Black RE. Patterns of growth in early childhood and infectious disease and nutritional determinants. Nestle Nutr Inst Workshop Ser 2017;87:63–72.

18. Dewey KG, Vitta BS. Strategies for ensuring adequate nutrient intake for infants and young children during the period of complementary feeding. Insight Issue No 7, pp 14 >Alive & Thrive Technical Brief: Washington DC 2013.

19. Arimond M, Zeilani M, Jungjohann S, Brown KH, Ashorn P, Allen LH, Dewey KG. Considerations in developing lipid-based nutrient supplements for prevention of undernutrition: experience from the International Lipid-Based Nutrient Supplements (iLiNS) Project. Matern Child Nutr 2015;11 Suppl 4:31–61.

20. Toure M, Becquey E, Huybregts L, Diatta D, Booth A, Verstraeten R. Evidence mapping of wasting programs and their impact along the continuum of care in low- and middle-income countries: a rapid review of the research evidence. Transform Nutrition West Africa Evidence Note 23 Dakar, Senegal: International Food Policy Research Institute, 2021; Available at: https://westafrica.transformnutrition.org/wp-content/uploads/2021/09/EvNote23_EvMappingWastingPrograms.pdf. Accessed on: 10 June 2022.

21. Maximizing the Quality of Scaling Up Nutrition Plus (MQSUN+). The current state of evidence and thinking on wasting prevention - final report. Washington, DC: MQSUN+, 2018. Available at: https://mqsunplus.path.org/resources/the-current-state-of-evidence-and-thinking-on-wasting-prevention/. Accessed on: 10 June 2022.

22. Dewey KG, Stewart CP, Wessells KR, Prado EL, Arnold CD. Small-quantity lipid-based nutrient supplements for the prevention of child malnutrition and promotion of healthy development: overview of individual participant data meta-analysis and programmatic implications. Am J Clin Nutr 2021;114:3S–14S.

23. Dewey KG, Wessells KR, Arnold CD, Prado EL, Abbeddou S, Adu-Afarwuah S, Ali H, Arnold BF, Ashorn P, Ashorn U, et al. Characteristics that modify the effect of small-quantity lipid-based nutrient supplementation on child growth: an individual participant data meta-analysis of randomized controlled trials. Am J Clin Nutr 2021;114:15S–42S.

24. Prado EL, Arnold CD, Wessells KR, Stewart CP, Abbeddou S, Adu-Afarwuah S, Arnold BF, Ashorn U, Ashorn P, Becquey E, et al. Small-quantity lipid-based nutrient supplements for children age 6-24 months: a systematic review and individual participant data meta-analysis of effects on developmental outcomes and effect modifiers. Am J Clin Nutr 2021;114:43S–67S.

25. Wessells KR, Arnold CD, Stewart CP, Prado EL, Abbeddou S, Adu-Afarwuah S, Arnold BF, Ashorn P, Ashorn U, Becquey E, et al. Characteristics that modify the effect of small-quantity lipid-based nutrient supplementation on child anemia and micronutrient status: an individual participant data meta-analysis of randomized controlled trials. Am J Clin Nutr 2021;114:68S–94S.

26. Das JK, Salam RA, Hadi YB, Sadiq Sheikh S, Bhutta AZ, Weise Prinzo Z, Bhutta ZA. Preventive lipid-based nutrient supplements given with complementary foods to infants and young children 6 to 23 months of age for health, nutrition, and developmental outcomes. Cochrane Database Syst Rev 2019;5:CD012611.

27. Wessells K, Dewey K, Stewart C, Arnold C, Prado E. Modifiers of the effect of LNS provided to infants and children 6 to 24 months of age on growth outcomes: a systematic review and meta-analysis of individual participant data from randomized controlled trials in low-income and middle-income countries. PROSPERO 2019 CRD42019146592 Available from: https://www.crd.york.ac.uk/prospero/display_record.php?ID=CRD42019146592.

28. Wessells KR, Stewart C, Arnold CD, Dewey K, Prado E. Modifiers of the effect of LNS provided to infants and children 6 to 24 months of age on growth, anemia, micronutrient status and development outcomes. Open Science Framework. Available from: https://osf.io/ymsfu.

29. Stewart LA, Clarke M, Rovers M, Riley RD, Simmonds M, Stewart G, Tierney JF, Group P-ID. Preferred reporting items for systematic review and meta-analyses of individual participant data: the PRISMA-IPD statement. JAMA 2015;313:1657–65.

30. World Bank Historical Classification by Income Group. Available at: http://databank.worldbank.org/data/download/site-content/OGHIST.xls Accessed on: 22 August 2019.

31. Johnston BC, Guyatt GH. Best (but oft-forgotten) practices: intention-to-treat, treatment adherence, and missing participant outcome data in the nutrition literature. Am J Clin Nutr 2016;104:1197–201.

32. World Health Organization Multicentre Growth Reference Study Group. WHO Child Growth Standards based on length/height, weight and age. Acta Paediatr Suppl 2006;450:76–85.

33. Tukey J. The future of data analysis. The annals of mathematical statistics 1962;33:1–67.

34. Higgins J, Green S. Cochrane Handbook for Systematic Reviews for Interventions, Version 5.1.0. The Cochrane Collaboration 2011. Available from www.handbook.cochrane.org.

35. Balshem H, Helfand M, Schunemann HJ, Oxman AD, Kunz R, Brozek J, Vist GE, Falck-Ytter Y, Meerpohl J, Norris S, et al. GRADE guidelines: 3. Rating the quality of evidence. J Clin Epidemiol 2011;64:401–6.

36. Burke DL, Ensor J, Riley RD. Meta-analysis using individual participant data: one-stage and two- stage approaches, and why they may differ. Stat Med 2017;36:855–75.

37. Veroniki AA, Jackson D, Viechtbauer W, Bender R, Bowden J, Knapp G, Kuss O, Higgins JP, Langan D, Salanti G. Methods to estimate the between-study variance and its uncertainty in meta- analysis. Res Synth Methods 2016;7:55–79.

38. Paule RC, Mandel J. Consensus values, regressions, and weighting factors. J Res Natl Inst Stand Technol 1989;94:197–203.

39. Higgins JP, Thompson SG, Deeks JJ, Altman DG. Measuring inconsistency in meta-analyses. BMJ 2003;327:557–60.

40. Streiner DL. Best (but oft-forgotten) practices: the multiple problems of multiplicity-whether and how to correct for many statistical tests. Am J Clin Nutr 2015;102:721–8.

41. Vancak V, Goldberg Y, Levine S. Systematic analysis of the number needed to treat. Statistical Methods in Medical Research 2020;29:2393–410.

42. Christian P, Shaikh S, Shamim AA, Mehra S, Wu L, Mitra M, Ali H, Merrill RD, Choudhury N, Parveen M, et al. Effect of fortified complementary food supplementation on child growth in rural Bangladesh: a cluster-randomized trial. Int J Epidemiol 2015;44:1862–76.

43. Dewey KG, Mridha MK, Matias SL, Arnold CD, Cummins JR, Khan MS, Maalouf-Manasseh Z, Siddiqui Z, Ullah MB, Vosti SA. Lipid-based nutrient supplementation in the first 1000 d improves child growth in Bangladesh: a cluster-randomized effectiveness trial. Am J Clin Nutr 2017;105:944–57.

44. Luby SP, Rahman M, Arnold BF, Unicomb L, Ashraf S, Winch PJ, Stewart CP, Begum F, Hussain F, Benjamin-Chung J, et al. Effects of water quality, sanitation, handwashing, and nutritional interventions on diarrhoea and child growth in rural Bangladesh: a cluster randomised controlled trial. Lancet Glob Health 2018;6:e302–e15.

45. Hess SY, Abbeddou S, Jimenez EY, Some JW, Vosti SA, Ouedraogo ZP, Guissou RM, Ouedraogo JB, Brown KH. Small-quantity lipid-based nutrient supplements, regardless of their zinc content, increase growth and reduce the prevalence of stunting and wasting in young Burkinabe children: a cluster-randomized trial. PLoS One 2015;10:e0122242.

46. Becquey E, Huybregts L, Zongrone A, Le Port A, Leroy JL, Rawat R, Toure M, Ruel MT. Impact on child acute malnutrition of integrating a preventive nutrition package into facility-based screening for acute malnutrition during well-baby consultation: a cluster-randomized controlled trial in Burkina Faso. PLoS Med 2019;16:e1002877.

47. Adu-Afarwuah S, Lartey A, Brown KH, Zlotkin S, Briend A, Dewey KG. Randomized comparison of 3 types of micronutrient supplements for home fortification of complementary foods in Ghana: effects on growth and motor development. Am J Clin Nutr 2007;86:412–20.

48. Adu-Afarwuah S, Lartey A, Okronipa H, Ashorn P, Peerson JM, Arimond M, Ashorn U, Zeilani M, Vosti S, Dewey KG. Small-quantity, lipid-based nutrient supplements provided to women during pregnancy and 6 mo postpartum and to their infants from 6 mo of age increase the mean attained length of 18-mo-old children in semi-urban Ghana: a randomized controlled trial. Am J Clin Nutr 2016;104:797–808.

49. Iannotti LL, Dulience SJ, Green J, Joseph S, Francois J, Antenor ML, Lesorogol C, Mounce J, Nickerson NM. Linear growth increased in young children in an urban slum of Haiti: a randomized controlled trial of a lipid-based nutrient supplement. Am J Clin Nutr 2014;99:198–208.

50. Null C, Stewart CP, Pickering AJ, Dentz HN, Arnold BF, Arnold CD, Benjamin-Chung J, Clasen T, Dewey KG, Fernald LCH, et al. Effects of water quality, sanitation, handwashing, and nutritional interventions on diarrhoea and child growth in rural Kenya: a cluster-randomised controlled trial. Lancet Glob Health 2018;6:e316–e29.

51. Galasso E, Weber AM, Stewart CP, Ratsifandrihamanana L, Fernald LCH. Effects of nutritional supplementation and home visiting on growth and development in young children in Madagascar: a cluster-randomised controlled trial. Lancet Glob Health 2019;7:e1257–e68.

52. Ashorn P, Alho L, Ashorn U, Cheung YB, Dewey KG, Gondwe A, Harjunmaa U, Lartey A, Phiri N, Phiri TE, et al. Supplementation of maternal diets during pregnancy and for 6 months postpartum and infant diets thereafter with small-quantity lipid-based nutrient supplements does not promote child growth by 18 months of age in rural Malawi: a randomized controlled trial. J Nutr 2015;145:1345–53.

53. Maleta KM, Phuka J, Alho L, Cheung YB, Dewey KG, Ashorn U, Phiri N, Phiri TE, Vosti SA, Zeilani M, et al. Provision of 10-40 g/d lipid-based nutrient supplements from 6 to 18 months of age does not prevent linear growth faltering in Malawi. J Nutr 2015;145:1909–15.

54. Huybregts L, Le Port A, Becquey E, Zongrone A, Barba FM, Rawat R, Leroy JL, Ruel MT. Impact on child acute malnutrition of integrating small-quantity lipid-based nutrient supplements into community-level screening for acute malnutrition: a cluster-randomized controlled trial in Mali. PLoS Med 2019;16:e1002892.

55. Humphrey JH, Mbuya MNN, Ntozini R, Moulton LH, Stoltzfus RJ, Tavengwa NV, Mutasa K, Majo F, Mutasa B, Mangwadu G, et al. Independent and combined effects of improved water, sanitation, and hygiene, and improved complementary feeding, on child stunting and anaemia in rural Zimbabwe: a cluster-randomised trial. Lancet Glob Health 2019;7:e132–e47.

56. Prendergast AJ, Chasekwa B, Evans C, Mutasa K, Mbuya MNN, Stoltzfus RJ, Smith LE, Majo FD, Tavengwa NV, Mutasa B, et al. Independent and combined effects of improved water, sanitation, and hygiene, and improved complementary feeding, on stunting and anaemia among HIV-exposed children in rural Zimbabwe: a cluster-randomised controlled trial. Lancet Child Adolesc Health 2019;3:77–90.

57. Smuts CM, Matsungo TM, Malan L, Kruger HS, Rothman M, Kvalsvig JD, Covic N, Joosten K, Osendarp SJM, Bruins MJ, et al. Effect of small-quantity lipid-based nutrient supplements on growth, psychomotor development, iron status, and morbidity among 6- to 12-mo-old infants in South Africa: a randomized controlled trial. Am J Clin Nutr 2019;109:55–68.

58. Stewart CP, Wessells KR, Arnold CD, Huybregts L, Ashorn P, Becquey E, Humphrey JH, Dewey KG. Lipid-based nutrient supplements and all-cause mortality in children 6-24 months of age: a meta-analysis of randomized controlled trials. Am J Clin Nutr 2020;111:207–18.

59. Adams K, Vosti S, Arnold C, Engle-Stone R, Prado E, Stewart C, Wessells K, Dewey K. The cost-effectiveness of small-quantity lipid-based nutrient supplements for prevention of child death and malnutrition and promotion of healthy development: modeling results for Uganda. medRxiv 2022052722275713; doi: https://doiorg/101101/2022052722275713 2022.

