## Supplemental Figures Table of Contents for "Preventive small-quantity lipid-based nutrient supplements reduce severe wasting and severe stunting among young children: an individual participant data meta-analysis of randomized controlled trials"

Dewey *et al.* (2022)

**Table of Contents: Supplemental Figures**

Supplemental Box 1: Potential effect modifiers

Supplemental Figure 1: Study flow diagram

Supplemental Figure 2: Summary risk of bias as a percentage of all included studies for the effects of SQ-LNS on growth outcomes

Supplemental Figure 3: Forest plot of effect of SQ-LNS on prevalence of severe acute malnutrition (SAM)

Supplemental Figure 4: Forest plot of effect of SQ-LNS on prevalence of very low MUAC

Supplemental Figure 5: Sensitivity analyses of main effects of SQ-LNS on prevalence ratios for severe acute malnutrition (SAM)

Supplemental Figure 6: Sensitivity analyses of main effects of SQ-LNS on prevalence ratios for very low MUAC

Supplemental Figure 7: Forest plot of effect of SQ-LNS on severe wasting prevalence using the rare events sensitivity analysis, with estimates generated for certain trials in which 0.5 is substituted for the 0 value in analysis

Supplemental Figures 8 A to I: Forest plots for severe wasting, stratified by study-level characteristics

Supplemental Figures 9 A to I: Forest plots for severe stunting, stratified by study-level characteristics
