## Supplemental Box 1 for "Preventive small-quantity lipid-based nutrient supplements reduce severe wasting and severe stunting among young children: an individual participant data meta-analysis of randomized controlled trials"

**Supplemental Box 1. Potential effect modifiers^1^**

| **Study-level effect modifiers** | **Individual-level maternal, child and household effect modifiers** |
| --- | --- |
| - Geographic region (WHO region: African vs. South-East Asia Region) - Stunting burden among control group children at 18 mo of age (≥ 35% vs. < 35%)^2^ - Wasting burden among control group children at endline (≥ 10% vs. < 10%) - Malaria prevalence (country-specific, closest in time to the study: ≥ 10% vs. < 10%)^3^ - Water quality (study-specific, < 75% vs. ≥ 75% prevalence of improved drinking water)^4^ - Sanitation (study-specific, < 50% vs. ≥ 50% prevalence of improved sanitation)^5^ - Duration of child supplementation (study target: >12 mo vs. ≤ 12 mo) - Child age at baseline or endline - Frequency of contact for intervention delivery or outcome assessments during the study (weekly vs monthly) - Compliance (average percent compliance in LNS group ≥ 80% vs. ≤ 80%)^6^ | - Maternal height (< 150.1 cm vs. ≥ 150.1 cm)^7^ - Maternal BMI (< 20 kg/m^2^ vs. ≥ 20 kg/m^2^) - Maternal age (< 25 y vs. ≥ 25 y) - Maternal education (no formal or incomplete primary vs. complete primary or greater) - Maternal depressive symptoms (lower, < study 75^th^ percentile vs. higher, ≥ study 75^th^ percentile)^8^ - Child sex (female vs. male) - Child birth order (first born vs. later born) - Child baseline anthropometric status (lower vs. higher)^9^ - Household socio-economic status (< study median vs. ≥ study median)^10^ - Food security (moderate to severe food insecurity vs. mild to secure)^11^ - Source water quality (unimproved vs. improved)^4^ - Sanitation (unimproved vs. improved)^4^ - Home environment (< study median vs. ≥ study median)^12^ - Season at the time of growth outcome assessment (rainy vs. dry)^13^ |

LNS, lipid-based nutrient supplements

^1^ Reference category on the right (non-reference vs. reference)

^2^Based on 18-mo data because baseline data not available for all trials; cutoff chosen at approximately the median across trials

^3^ World Malaria Report 2018 (1); cutoff chosen based on the median across trials

^4^ improved water source includes piped water, boreholes or tubewells, protected dug wells or springs, rainwater, packaged or delivered water, see Supplemental Table 2 (2); based on baseline data, excluding arms that received WASH interventions; cutoff chosen at approximately the median across trials

^5^ improved sanitation includes flush/pour flush to piped sewer system, septic tanks or pit latrines; ventilated improved pit latrines, composting toilets or pit latrines with slabs, see Supplemental Table 2 (3); based on baseline data, excluding arms that received WASH interventions; cutoff chosen at approximately the median across trials

^6^ study-specific, as reported based on a study-defined indicator (see Supplemental Table 2); cutoff chosen based on the median across trials

^7^ cutoff is -2 SD for height at 19 years of age: <https://www.who.int/growthref/hfa_girls_5_19years_z.pdf?ua=1>

^8^ study-specific; cutoff chosen to reflect top quartile for risk of depression

^9^ LAZ < vs. ≥ -1 when stunting is the outcome; WLZ < vs. ≥ 0 wasting or acute malnutrition is the outcome; MUACZ < vs. ≥ 0 when low MUAC is the outcome; MUAC, mid-upper arm circumference; MUACZ, mid-upper arm circumference z-score; LAZ, length-for-age z-score; WLZ, weight-for-length z-score.

^10^ based on a study-defined, study-specific assets index

^11^ study-specific (4)

^12^ as measured by the Family Care Indicators, Home Observation for the Measurement of the Environment Inventory, or other similar tools (4)

^13^ rainy vs. dry, based on study- and child-specific average rainfall during the month of measurement and two months prior (4)

**References**

1.	World Malaria Report 2018. Geneva: World Health Organization; 2018. Available at: <https://www.who.int/malaria/publications/world-malaria-report-2018/report/en/> Accessed on: 26 August 2019.

2.	WHO, UNICEF Joint Monitoring Programme. Drinking Water. Available at <http://washdata.org/monitoring/drinking-water>. Accessed on: 26 August 2019.

3.	WHO, UNICEF Joint Monitoring Programme. Sanitation. Available at <http://washdata.org/monitoring/sanitation>. Accessed on: 26 August 2019.

4.	Dewey KG, Wessells KR, Arnold CD, Prado EL, Abbeddou S, Adu-Afarwuah S, Ali H, Arnold BF, Ashorn P, Ashorn U, et al. Characteristics that modify the effect of small-quantity lipid-based nutrient supplementation on child growth: an individual participant data meta-analysis of randomized controlled trials. Am J Clin Nutr 2021;114:15S-42S.
