## Supplemental Figure 1 for "Preventive small-quantity lipid-based nutrient supplements reduce severe wasting and severe stunting among young children: an individual participant data meta-analysis of randomized controlled trials"

Supplemental Figure 1: Study flow diagram

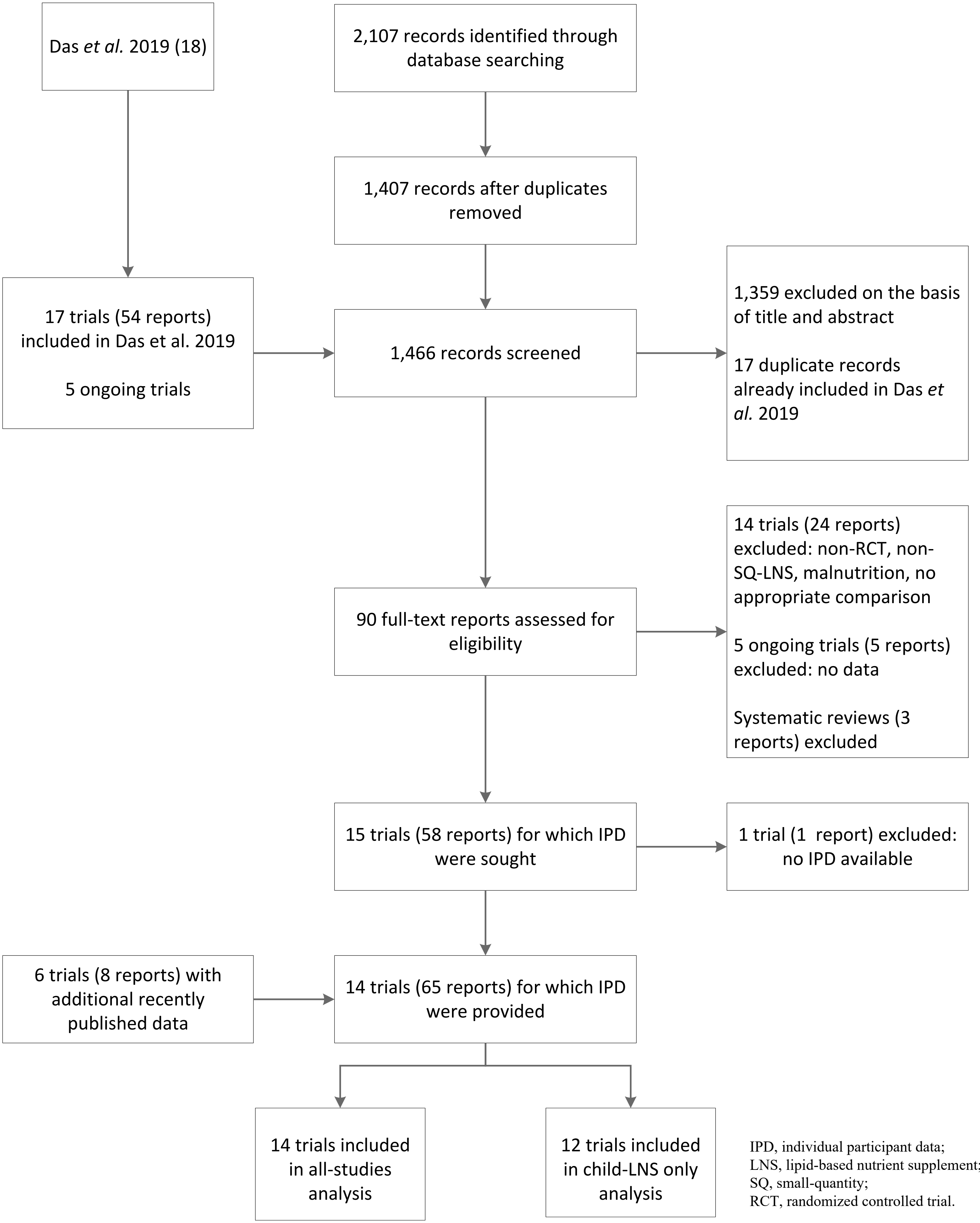


IPD, individual participant data;
LNS, lipid-based nutrient supplement;
SQ, small-quantity;
RCT, randomized controlled trial.
