## Supplemental Figure 2 for "Preventive small-quantity lipid-based nutrient supplements reduce severe wasting and severe stunting among young children: an individual participant data meta-analysis of randomized controlled trials"

Supplemental Figure 2: Summary risk of bias as a percentage of all included studies for the effects of SQ-LNS on growth outcomes

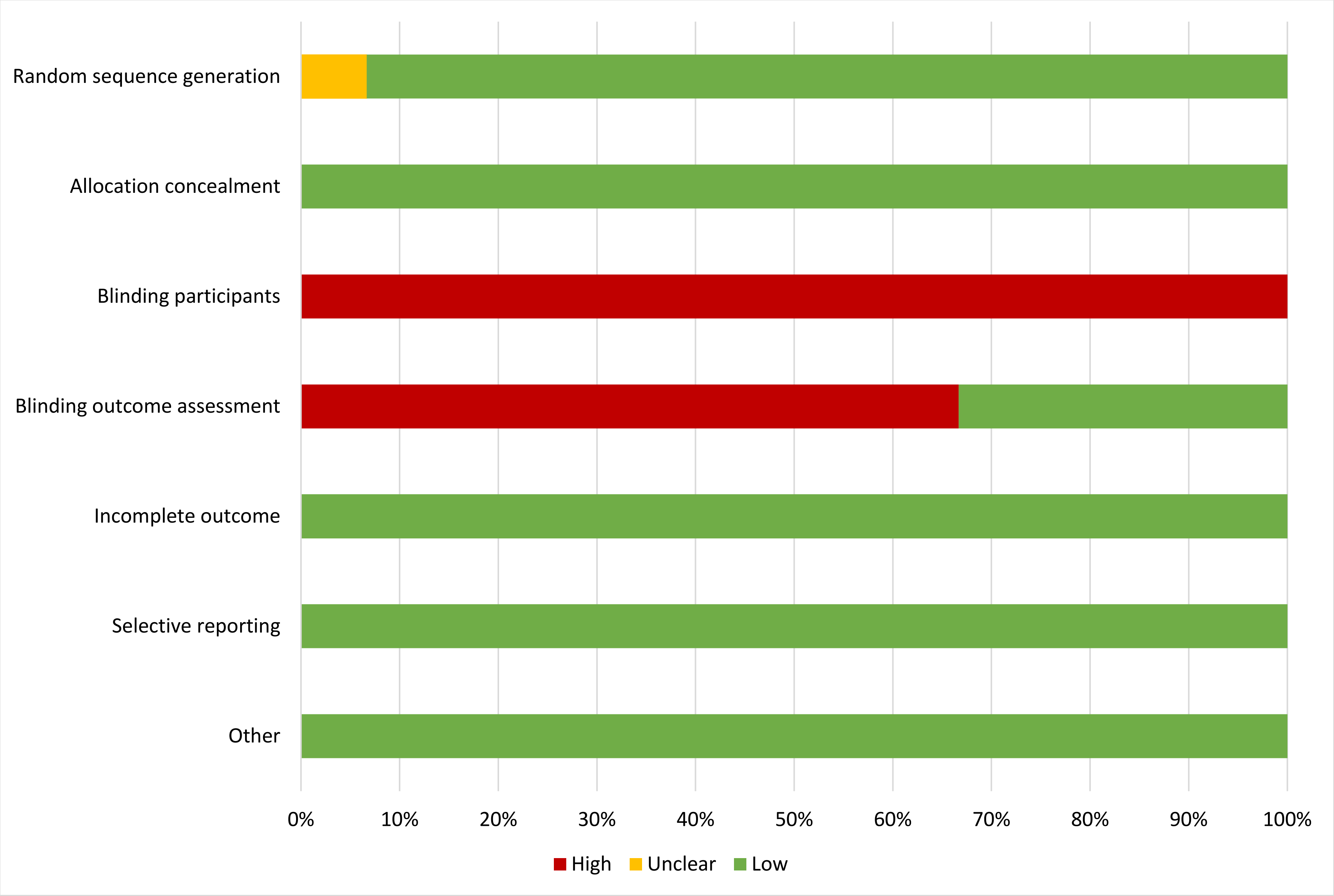
