## Supplemental Figure 3 for "Preventive small-quantity lipid-based nutrient supplements reduce severe wasting and severe stunting among young children: an individual participant data meta-analysis of randomized controlled trials"

Supplemental Figure 3: Forest plot of effect of SQ-LNS on prevalence of severe acute malnutrition (SAM)

| Country | Trial | LNS N | Control N | PR (95% CI) | Fixed W | Random W |
|---|---|---|---|---|---|---|
| Bangladesh | JiVitA-4 (42) | 2783 | 1216 | 0.78 (0.52, 1.16) | 0.26 | 0.26 |
| Bangladesh | RDNS (43) | 1661 | 815 | 0.70 (0.34, 1.45) | 0.08 | 0.08 |
| Bangladesh | WASH-B (44) | | | | | |
| Burkina Faso | iLiNS-Zinc (45) | 1951 | 664 | 0.39 (0.21, 0.75) | 0.10 | 0.10 |
| Burkina Faso | PROMIS (46) | 846 | 901 | 1.33 (0.49, 3.62) | 0.04 | 0.04 |
| Burkina Faso | PROMIS (46) | 424 | 434 | 0.77 (0.40, 1.47) | 0.10 | 0.10 |
| Ghana | GHANA (47) | | | | | |
| Ghana | iLiNS-DYAD-G (48) | 347 | 692 | 0.85 (0.22, 3.28) | 0.02 | 0.02 |
| Haiti | HAITI (49) | | | | | |
| Kenya | WASH-B (50) | 1454 | 5117 | 0.67 (0.27, 1.69) | 0.05 | 0.05 |
| Madagascar | MAHAY (51) | 1675 | 1655 | 0.81 (0.47, 1.39) | 0.14 | 0.14 |
| Malawi | iLiNS-DYAD-M (52) | 220 | 444 | 1.01 (0.25, 4.00) | 0.02 | 0.02 |
| Malawi | iLiNS-DOSE (53) | 690 | 240 | 2.34 (0.53, 10.33) | 0.02 | 0.02 |
| Mali | PROMIS (54) | | | | | |
| Mali | PROMIS (54) | 944 | 959 | 0.76 (0.38, 1.53) | 0.09 | 0.09 |
| Zimbabwe | SHINE (HIV-) (55) | 1866 | 1775 | 0.86 (0.37, 1.97) | 0.06 | 0.06 |
| Zimbabwe | SHINE (HIV+) (56) | 336 | 327 | 0.65 (0.11, 3.69) | 0.01 | 0.01 |
| | **$I^2$ = 0.00, $Tau^2$ = 0.00** | **15197** | **15239** | | | |
| | **Fixed** | | | **0.76 (0.62, 0.93)** | | |
| | **Random** | | | **0.76 (0.62, 0.93)** | | |

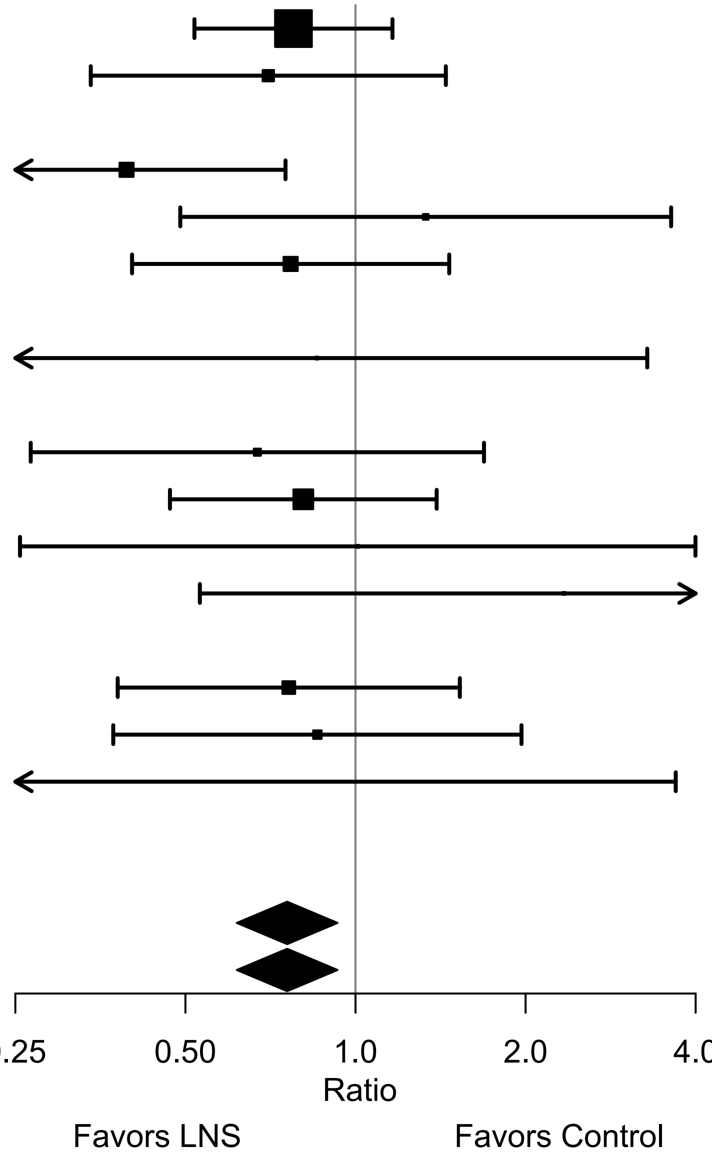


LNS, lipid-based nutrient supplements; PR, prevalence ratio.
Individual study estimates were generated from log-binomial regression controlling for baseline measure when available and with clustered observations using robust standard errors for cluster-randomized trials. Pooled estimates were generated using inverse-variance weighting with both fixed and random effects.
