## Supplemental Figure 4 for "Preventive small-quantity lipid-based nutrient supplements reduce severe wasting and severe stunting among young children: an individual participant data meta-analysis of randomized controlled trials"

Supplemental Figure 4: Forest plot of effect of SQ-LNS on prevalence of very low MUAC

| Country | Trial | LNS N | Control N | PR (95% CI) | Fixed W | Random W |
|---|---|---|---|---|---|---|
| Bangladesh | JiVitA-4 (42) | 2897 | 1271 | 0.90 (0.47, 1.73) | 0.16 | 0.15 |
| Bangladesh | RDNS (43) | 1663 | 815 | 2.45 (0.29, 20.42) | 0.01 | 0.02 |
| Bangladesh | WASH-B (44) | | | | | |
| Burkina Faso | iLiNS-Zinc (45) | 1959 | 666 | 0.38 (0.22, 0.65) | 0.23 | 0.19 |
| Burkina Faso | PROMIS (46) | 854 | 912 | 1.50 (0.57, 3.93) | 0.07 | 0.08 |
| Burkina Faso | PROMIS (46) | 424 | 436 | 1.03 (0.42, 2.50) | 0.08 | 0.09 |
| Ghana | GHANA (47) | | | | | |
| Ghana | iLiNS-DYAD-G (48) | | | | | |
| Haiti | HAITI (49) | | | | | |
| Kenya | WASH-B (50) | 1461 | 5185 | 0.39 (0.17, 0.93) | 0.09 | 0.10 |
| Madagascar | MAHAY (51) | 1680 | 1658 | 1.03 (0.54, 1.95) | 0.16 | 0.15 |
| Malawi | iLiNS-DYAD-M (52) | 220 | 444 | 0.81 (0.16, 4.13) | 0.03 | 0.03 |
| Malawi | iLiNS-DOSE (53) | 696 | 240 | 0.51 (0.09, 3.03) | 0.02 | 0.03 |
| Mali | PROMIS (54) | 507 | 505 | 1.00 (0.06, 15.67) | 0.01 | 0.01 |
| Mali | PROMIS (54) | 954 | 970 | 0.95 (0.44, 2.06) | 0.11 | 0.11 |
| Zimbabwe | SHINE (HIV-) (55) | 1872 | 1780 | 0.38 (0.07, 1.94) | 0.03 | 0.03 |
| Zimbabwe | SHINE (HIV+) (56) | | | | | |
| | **I² = 0.24, Tau² = 0.04** | **15187** | **14882** | | | |
| | **Fixed** | | | **0.73 (0.56, 0.94)** | | |
| | **Random** | | | **0.74 (0.55, 1.00)** | | |

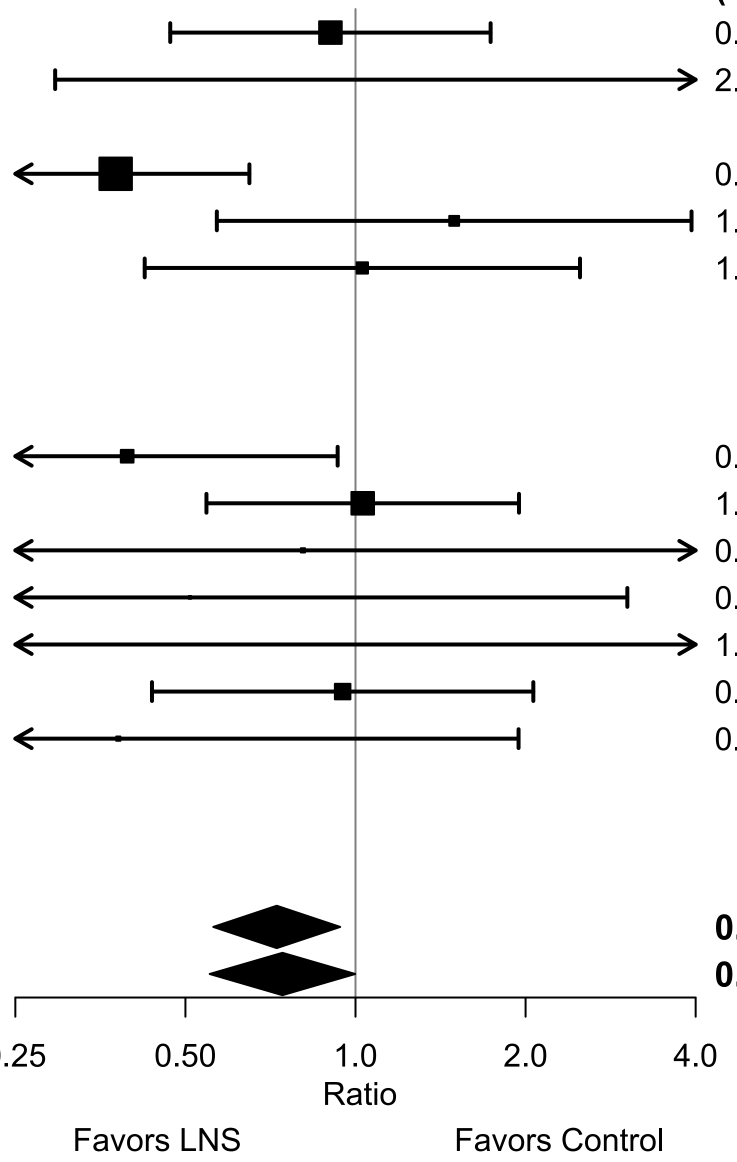


LNS, lipid-based nutrient supplements; MUAC, mid-upper arm circumference; PR, prevalence ratio.
Individual study estimates were generated from log-binomial regression controlling for baseline measure when available and with clustered observations using robust standard errors for cluster-randomized trials. Pooled estimates were generated using inverse-variance weighting with both fixed and random effects.
