## Supplemental Figure 5 for "Preventive small-quantity lipid-based nutrient supplements reduce severe wasting and severe stunting among young children: an individual participant data meta-analysis of randomized controlled trials"

Supplemental Figure 5: Sensitivity analyses of main effects of SQ-LNS on prevalence ratios for severe acute malnutrition (SAM)

| **Analysis** | **LNS N** | **Control N** | **Pooled Estimate (95% CI)** |
|---|---|---|---|
| All-trials | 15197 | 15239 | 0.76 (0.62, 0.93) |
| Zero event trials included | 15701 | 15745 | 0.76 (0.62, 0.93) |
| Child-LNS-only | 12938 | 14103 | 0.77 (0.62, 0.96) |
| Multi-component | 12602 | 11619 | 0.78 (0.62, 0.96) |
| Passive arms excluded | 10987 | 12724 | 0.84 (0.67, 1.05) |
| Milk-peanut-LNS only | 11221 | 14103 | 0.79 (0.63, 1.00) |

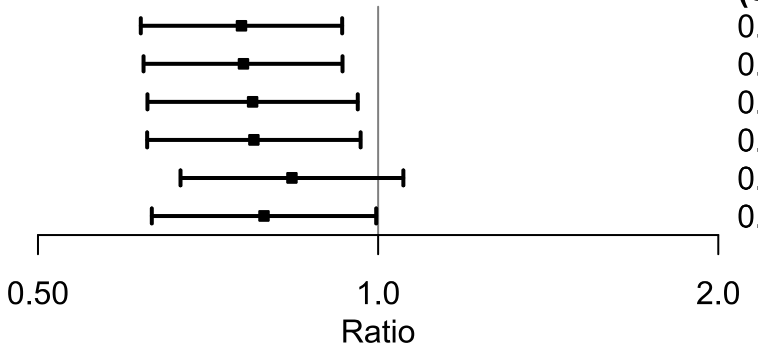


All-trial analysis includes all trials;
Child-LNS-only excludes trial arms that provided both maternal and child LNS;
Multi-component analysis separates comparisons within trials that included multi-component interventions, so that the SQ-LNS vs. no SQ-LNS comparisons were conducted separately between pairs of arms that included the same non-nutrition components (e.g. SQ-LNS+WASH vs. WASH; SQ-LNS vs. Control);
Passive arms excluded analysis excludes passive control arms;
Milk-peanut LNS only analysis excludes arms with SQ-LNS formulations that were not milk and peanut based;
“Zero events trials included” uses estimates generated for certain trials in which 0.5 is substituted for the 0 value in analysis for severe wasting.
LNS, lipid-based nutrient supplements.
