## Supplemental Figure 6 for "Preventive small-quantity lipid-based nutrient supplements reduce severe wasting and severe stunting among young children: an individual participant data meta-analysis of randomized controlled trials"

Supplemental Figure 6: Sensitivity analyses of main effects of SQ-LNS on prevalence ratios for very low MUAC

| Analysis | LNS N | Control N | Pooled Estimate (95% CI) |
|---|---|---|---|
| All-trials | 15187 | 14882 | 0.73 (0.56, 0.94) |
| Zero event trials included | 15871 | 15902 | 0.73 (0.56, 0.94) |
| Child-LNS-only | 13272 | 14438 | 0.73 (0.56, 0.96) |
| Multi-component | 13272 | 12255 | 0.75 (0.57, 0.98) |
| Passive arms excluded | 11313 | 13050 | 0.90 (0.66, 1.22) |
| Milk-peanut-LNS only | 11499 | 14438 | 0.74 (0.56, 0.97) |

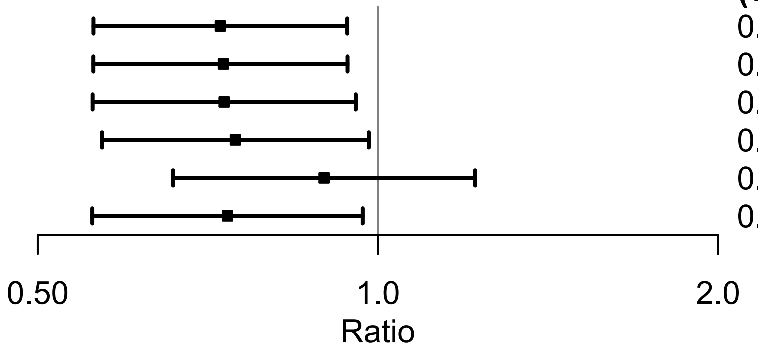


All-trial analysis includes all trials;
Child-LNS-only excludes trial arms that provided both maternal and child LNS;
Multi-component analysis separates comparisons within trials that included multi-component interventions, so that the SQ-LNS vs. no SQ-LNS comparisons were conducted separately between pairs of arms that included the same non-nutrition components (e.g. SQ-LNS+WASH vs. WASH; SQ-LNS vs. Control);
Passive arms excluded analysis excludes passive control arms;
Milk-peanut LNS only analysis excludes arms with SQ-LNS formulations that were not milk and peanut based;
“Zero events trials included” uses estimates generated for certain trials in which 0.5 is substituted for the 0 value in analysis for severe wasting.
LNS, lipid-based nutrient supplements; MUAC, mid-upper arm circumference.
