## Supplemental Figure 7 for "Preventive small-quantity lipid-based nutrient supplements reduce severe wasting and severe stunting among young children: an individual participant data meta-analysis of randomized controlled trials"

Supplemental Figure 7: Forest plot of effect of SQ-LNS on severe wasting prevalence using the rare events sensitivity analysis, with estimates generated for certain trials in which 0.5 is substituted for the 0 value in analysis

| Country | Trial | LNS N | Control N | PR (95% CI) | Fixed W | Random W |
|---|---|---|---|---|---|---|
| Bangladesh | JiVitA-4 (42) | 2783 | 1217 | 0.90 (0.58, 1.39) | 0.27 | 0.22 |
| Bangladesh | RDNS (43) | 1661 | 815 | 0.70 (0.34, 1.45) | 0.10 | 0.10 |
| Bangladesh | WASH-B (44) | 1156 | 3424 | 0.61 (0.30, 1.25) | 0.10 | 0.10 |
| Burkina Faso | iLiNS-Zinc (45) | 1952 | 664 | 0.36 (0.19, 0.68) | 0.12 | 0.12 |
| Burkina Faso | PROMIS (46) | 856 | 907 | 1.06 (0.33, 3.38) | 0.04 | 0.04 |
| Burkina Faso | PROMIS (46) | 430 | 436 | 0.51 (0.17, 1.54) | 0.04 | 0.05 |
| Ghana | GHANA (47) | 98 | 96 | 3.92 (0.45, 34.42) | 0.01 | 0.01 |
| Ghana | iLiNS-DYAD-G (48) | 347 | 692 | 0.85 (0.22, 3.28) | 0.03 | 0.03 |
| Haiti | HAITI (49) | 149 | 149 | 1.00 (0.02, 50.07) | 0.00 | 0.00 |
| Kenya | WASH-B (50) | 1455 | 5118 | 0.27 (0.04, 1.86) | 0.01 | 0.02 |
| Madagascar | MAHAY (51) | 1700 | 1682 | 0.58 (0.29, 1.17) | 0.10 | 0.10 |
| Malawi | iLiNS-DYAD-M (52) | 220 | 444 | 2.02 (0.41, 9.92) | 0.02 | 0.02 |
| Malawi | iLiNS-DOSE (53) | 696 | 241 | 7.62 (0.45, 129.13) | 0.01 | 0.01 |
| Mali | PROMIS (54) | 504 | 506 | 2.01 (0.07, 59.72) | 0.00 | 0.01 |
| Mali | PROMIS (54) | 944 | 959 | 0.48 (0.21, 1.10) | 0.08 | 0.08 |
| Zimbabwe | SHINE (HIV-) (55) | 1869 | 1784 | 0.95 (0.38, 2.38) | 0.06 | 0.07 |
| Zimbabwe | SHINE (HIV+) (56) | 336 | 328 | 0.98 (0.14, 6.63) | 0.01 | 0.02 |
| | **$I^2$ = 0.08, $Tau^2$ = 0.02** | **17156** | **19462** | | | |
| | **Fixed** | | | **0.70 (0.56, 0.88)** | | |
| | **Random** | | | **0.70 (0.55, 0.89)** | | |

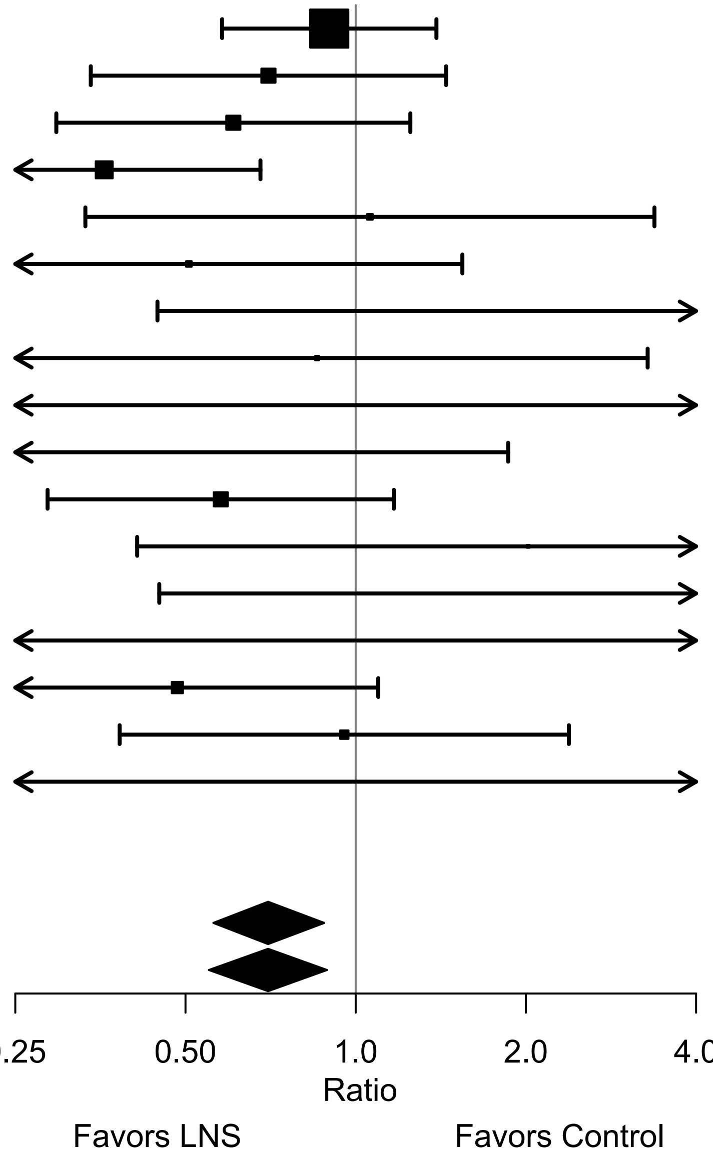


LNS, lipid-based nutrient supplements; PR, prevalence ratio.
Individual study estimates were generated from log-binomial regression controlling for baseline measure when available and with clustered observations using robust standard errors for cluster-randomized trials. Pooled estimates were generated using inverse-variance weighting with both fixed and random effects.
