## Supplemental Figure 8 for "Preventive small-quantity lipid-based nutrient supplements reduce severe wasting and severe stunting among young children: an individual participant data meta-analysis of randomized controlled trials"

### Supplemental figure 8: Forest plots for effects of SQ-LNS on severe wasting stratified by study-level effect modifiers

These figures are forest plots showing the study-level effect modification of intervention effects. Each figure shows the study-level estimates along with the corresponding pooled estimate grouped by study-level effect modifier category. Individual study estimates were generated from log-binomial regression; controlling for baseline measure when available and with clustered observations using robust standard errors for cluster-randomized trials. Pooled sub-group estimates were generated using inverse-variance weighting random effects. P-value for the difference was estimated using random effects meta-regression with the indicated effect modifier as the predictor of intervention effect size; stratified pooled estimates are presented for each strata.

The labels on the left y-axis correspond to trial level information. The values on the right indicate the study level effect estimate, confidence interval, and weighting for deriving the pooled estimates.

### Supplemental figure 8: Severe wasting prevalence ratio

#### 8A: Stratified by Geographic region

**Geographic region**
**(p−diff = 0.276)**

| **Geographic region − SEAR** | | | | **PR** | |
|---|---|---|---|---|---|
| **Country** | **Trial** | **N** | **N** | **(95% CI)** | **W** |
| Bangladesh | JiVitA−4 (42) | 2783 | 1217 | 0.90 (0.58, 1.39) | 0.57 |
| Bangladesh | RDNS (43) | 1661 | 815 | 0.70 (0.34, 1.45) | 0.21 |
| Bangladesh | WASH−B (44) | 1156 | 3424 | 0.61 (0.30, 1.25) | 0.21 |
| **$I^2$ = 0.00, $Tau^2$ = 0.00** | | **5600** | **5456** | **0.78 (0.56, 1.09)** | |
| **Geographic region − AFR** | | | | | |
| Burkina Faso | iLiNS−Zinc (45) | 1952 | 664 | 0.36 (0.19, 0.68) | 0.24 |
| Burkina Faso | PROMIS (46) | 856 | 907 | 1.06 (0.33, 3.38) | 0.07 |
| Burkina Faso | PROMIS CS (46) | 430 | 436 | 0.51 (0.17, 1.54) | 0.08 |
| Ghana | GHANA (47) | 98 | 96 | 3.92 (0.45, 34.42) | 0.02 |
| Ghana | iLiNS−DYAD−G (48) | 347 | 692 | 0.85 (0.22, 3.28) | 0.05 |
| Kenya | WASH−B (50) | 1455 | 5118 | 0.27 (0.04, 1.86) | 0.03 |
| Madagascar | MAHAY (51) | 1700 | 1682 | 0.58 (0.29, 1.17) | 0.19 |
| Malawi | iLiNS−DYAD−M (52) | 220 | 444 | 2.02 (0.41, 9.92) | 0.04 |
| Malawi | iLiNS−DOSE (53) | | | | |
| Mali | PROMIS (54) | | | | |
| Mali | PROMIS CS (54) | 944 | 959 | 0.48 (0.21, 1.10) | 0.14 |
| Zimbabwe | SHINE (HIV−) (55) | 1869 | 1784 | 0.95 (0.38, 2.38) | 0.11 |
| Zimbabwe | SHINE (HIV+) (56) | 336 | 328 | 0.98 (0.14, 6.63) | 0.03 |
| **$I^2$ = 0.09, $Tau^2$ = 0.03** | | **10207** | **13110** | **0.63 (0.45, 0.87)** | |

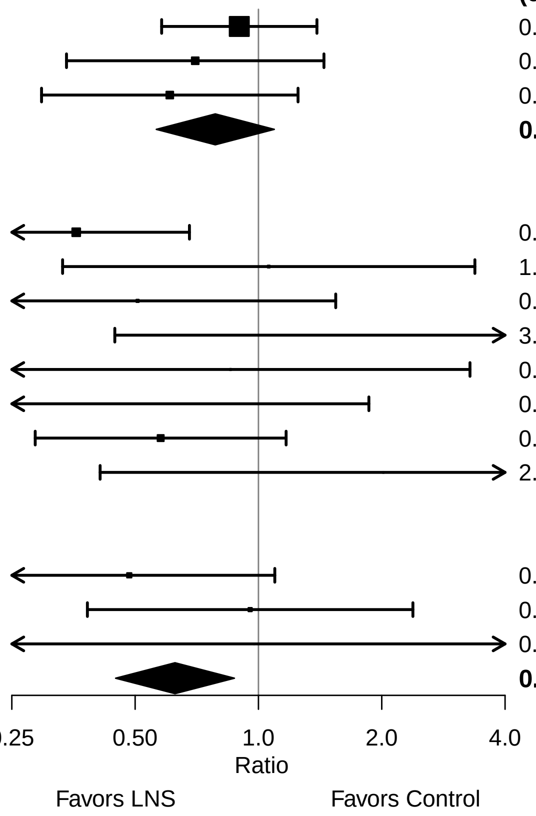

Supplemental figure 8: Severe wasting prevalence ratio

#### 8B: Stratified by Stunting burden

**Stunting burden**
**(p−diff = 0.343)**

| Country | Trial | N | N | PR (95% CI) | W |
|---|---|---|---|---|---|
| **Stunting burden − Less than 35%** | | | | | |
| Burkina Faso | PROMIS (46) | 856 | 907 | 1.06 (0.33, 3.38) | 0.25 |
| Burkina Faso | PROMIS CS (46) | 430 | 436 | 0.51 (0.17, 1.54) | 0.27 |
| Ghana | GHANA (47) | 98 | 96 | 3.92 (0.45, 34.42) | 0.07 |
| Ghana | iLiNS−DYAD−G (48) | 347 | 692 | 0.85 (0.22, 3.28) | 0.19 |
| Haiti | HAITI (49) | | | | |
| Kenya | WASH−B (50) | 1455 | 5118 | 0.27 (0.04, 1.86) | 0.09 |
| Malawi | iLiNS−DYAD−M (52) | 220 | 444 | 2.02 (0.41, 9.92) | 0.13 |
| **I² = 0.06, Tau² = 0.05** | | **3406** | **7693** | **0.89 (0.48, 1.64)** | |
| **Stunting burden − More than 35%** | | | | | |
| Bangladesh | JiVitA−4 (42) | 2783 | 1217 | 0.90 (0.58, 1.39) | 0.31 |
| Bangladesh | RDNS (43) | 1661 | 815 | 0.70 (0.34, 1.45) | 0.12 |
| Bangladesh | WASH−B (44) | 1156 | 3424 | 0.61 (0.30, 1.25) | 0.12 |
| Burkina Faso | iLiNS−Zinc (45) | 1952 | 664 | 0.36 (0.19, 0.68) | 0.15 |
| Madagascar | MAHAY (51) | 1700 | 1682 | 0.58 (0.29, 1.17) | 0.12 |
| Malawi | iLiNS−DOSE (53) | | | | |
| Mali | PROMIS (54) | | | | |
| Mali | PROMIS CS (54) | 944 | 959 | 0.48 (0.21, 1.10) | 0.09 |
| Zimbabwe | SHINE (HIV−) (55) | 1869 | 1784 | 0.95 (0.38, 2.38) | 0.07 |
| Zimbabwe | SHINE (HIV+) (56) | 336 | 328 | 0.98 (0.14, 6.63) | 0.02 |
| **I² = 0.00, Tau² = 0.00** | | **12401** | **10873** | **0.66 (0.51, 0.84)** | |

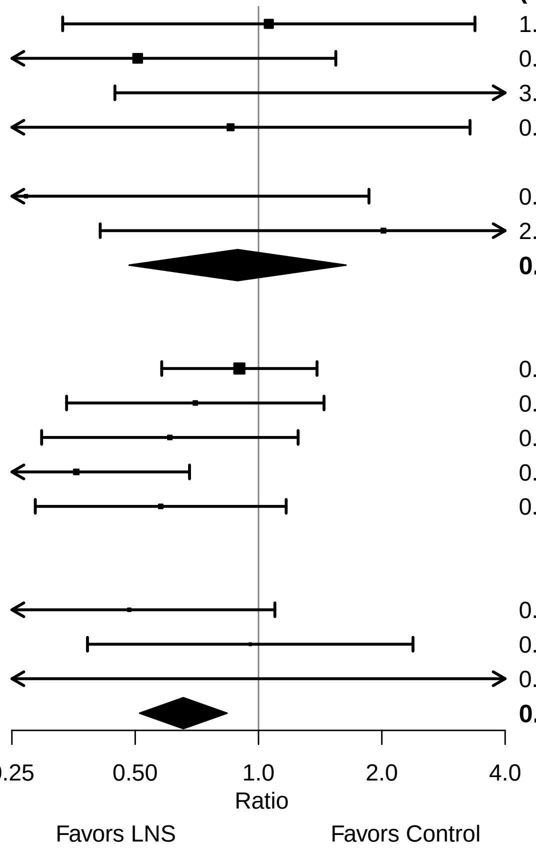

**Supplemental figure 8: Severe wasting prevalence ratio**

#### 8C: Stratified by Wasting burden

**Wasting burden**
**(p−diff = 0.354)**

**Wasting burden − Wasting < 10%**

| Country | Trial | N | N | PR (95% CI) | W |
|---|---|---|---|---|---|
| Ghana | GHANA (47) | 98 | 96 | 3.92 (0.45, 34.42) | 0.04 |
| Ghana | iLiNS−DYAD−G (48) | 347 | 692 | 0.85 (0.22, 3.28) | 0.11 |
| Haiti | HAITI (49) | | | | |
| Kenya | WASH−B (50) | 1455 | 5118 | 0.27 (0.04, 1.86) | 0.06 |
| Madagascar | MAHAY (51) | 1700 | 1682 | 0.58 (0.29, 1.17) | 0.41 |
| Malawi | iLiNS−DYAD−M (52) | 220 | 444 | 2.02 (0.41, 9.92) | 0.08 |
| Malawi | iLiNS−DOSE (53) | | | | |
| Zimbabwe | SHINE (HIV−) (55) | 1869 | 1784 | 0.95 (0.38, 2.38) | 0.24 |
| Zimbabwe | SHINE (HIV+) (56) | 336 | 328 | 0.98 (0.14, 6.63) | 0.06 |
| **I² = 0.00, Tau² = 0.00** | | **6025** | **10144** | **0.81 (0.52, 1.27)** | |

**Wasting burden − Wasting >= 10%**

| Country | Trial | N | N | PR (95% CI) | W |
|---|---|---|---|---|---|
| Bangladesh | JiVitA−4 (42) | 2783 | 1217 | 0.90 (0.58, 1.39) | 0.35 |
| Bangladesh | RDNS (43) | 1661 | 815 | 0.70 (0.34, 1.45) | 0.13 |
| Bangladesh | WASH−B (44) | 1156 | 3424 | 0.61 (0.30, 1.25) | 0.13 |
| Burkina Faso | iLiNS−Zinc (45) | 1952 | 664 | 0.36 (0.19, 0.68) | 0.17 |
| Burkina Faso | PROMIS (46) | 856 | 907 | 1.06 (0.33, 3.38) | 0.05 |
| Burkina Faso | PROMIS CS (46) | 430 | 436 | 0.51 (0.17, 1.54) | 0.06 |
| Mali | PROMIS (54) | | | | |
| Mali | PROMIS CS (54) | 944 | 959 | 0.48 (0.21, 1.10) | 0.10 |
| **I² = 0.13, Tau² = 0.01** | | **9782** | **8422** | **0.64 (0.48, 0.85)** | |

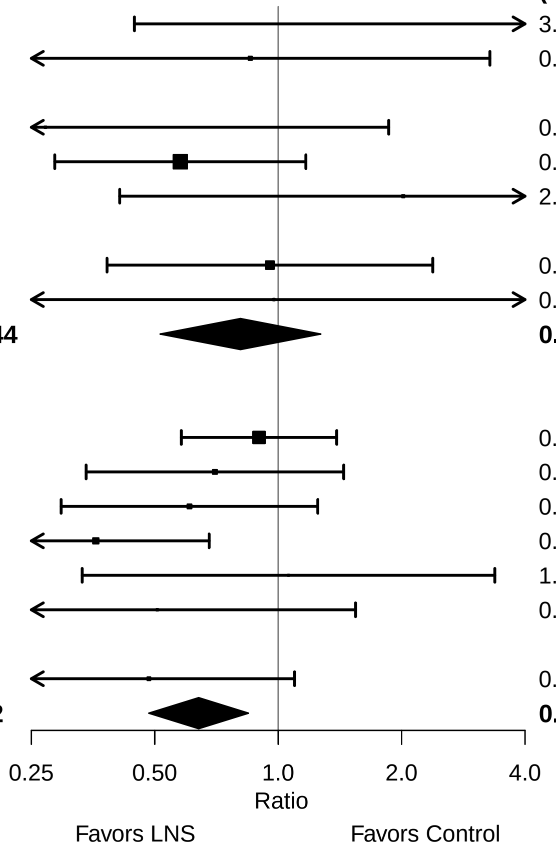

**Supplemental figure 8: Severe wasting prevalence ratio**

#### 8D: Stratified by Malaria prevalence

**Malaria prevalence**
**(p−diff = 0.294)**

| Country | Trial | N | N | PR (95% CI) | W |
|---|---|---|---|---|---|
| **Malaria prevalence − Less than 10%** | | | | | |
| Bangladesh | JiVitA−4 (42) | 2783 | 1217 | 0.90 (0.58, 1.39) | 0.40 |
| Bangladesh | RDNS (43) | 1661 | 815 | 0.70 (0.34, 1.45) | 0.15 |
| Bangladesh | WASH−B (44) | 1156 | 3424 | 0.61 (0.30, 1.25) | 0.15 |
| Haiti | HAITI (49) | | | | |
| Kenya | WASH−B (50) | 1455 | 5118 | 0.27 (0.04, 1.86) | 0.02 |
| Madagascar | MAHAY (51) | 1700 | 1682 | 0.58 (0.29, 1.17) | 0.16 |
| Zimbabwe | SHINE (HIV−) (55) | 1869 | 1784 | 0.95 (0.38, 2.38) | 0.09 |
| Zimbabwe | SHINE (HIV+) (56) | 336 | 328 | 0.98 (0.14, 6.63) | 0.02 |
| **I² = 0.00, Tau² = 0.00** | | **10960** | **14368** | **0.75 (0.57, 0.99)** | |
| **Malaria prevalence − At least 10%** | | | | | |
| Burkina Faso | iLiNS−Zinc (45) | 1952 | 664 | 0.36 (0.19, 0.68) | 0.37 |
| Burkina Faso | PROMIS (46) | 856 | 907 | 1.06 (0.33, 3.38) | 0.11 |
| Burkina Faso | PROMIS CS (46) | 430 | 436 | 0.51 (0.17, 1.54) | 0.12 |
| Ghana | GHANA (47) | 98 | 96 | 3.92 (0.45, 34.42) | 0.03 |
| Ghana | iLiNS−DYAD−G (48) | 347 | 692 | 0.85 (0.22, 3.28) | 0.08 |
| Malawi | iLiNS−DYAD−M (52) | 220 | 444 | 2.02 (0.41, 9.92) | 0.06 |
| Malawi | iLiNS−DOSE (53) | | | | |
| Mali | PROMIS (54) | | | | |
| Mali | PROMIS CS (54) | 944 | 959 | 0.48 (0.21, 1.10) | 0.22 |
| **I² = 0.34, Tau² = 0.18** | | **4847** | **4198** | **0.68 (0.40, 1.15)** | |

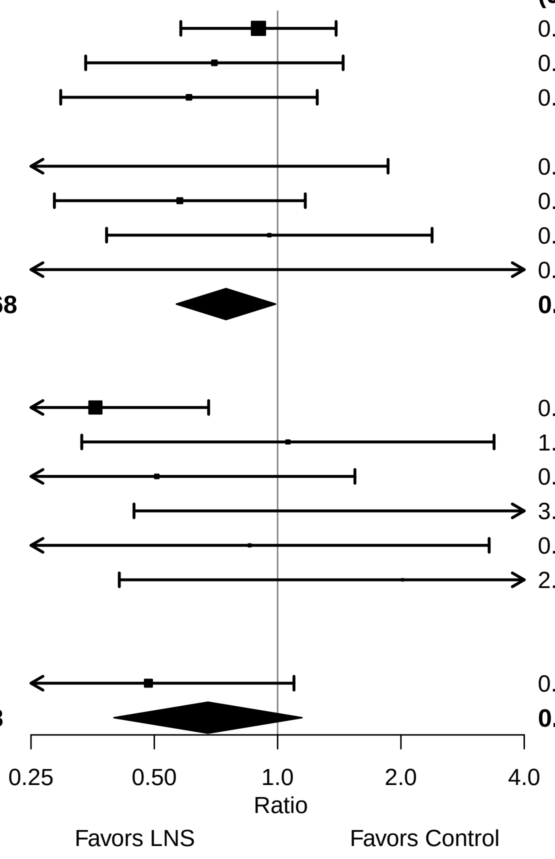

**Supplemental figure 8: Severe wasting prevalence ratio**

#### 8E: Stratified by Source water quality

**Source water quality**
**(p−diff = 0.035)**

| Country | Trial | N | N | PR (95% CI) | W |
|---|---|---|---|---|---|
| **Source water quality − Improved** | | | | | |
| Bangladesh | JiVitA−4 (42) | 2783 | 1217 | 0.90 (0.58, 1.39) | 0.57 |
| Bangladesh | RDNS (43) | 1661 | 815 | 0.70 (0.34, 1.45) | 0.22 |
| Bangladesh | WASH−B (44) | 566 | 1101 | 0.45 (0.14, 1.48) | 0.08 |
| Ghana | GHANA (47) | 98 | 96 | 3.92 (0.45, 34.42) | 0.02 |
| Ghana | iLiNS−DYAD−G (48) | 347 | 692 | 0.85 (0.22, 3.28) | 0.06 |
| Haiti | HAITI (49) | | | | |
| Malawi | iLiNS−DYAD−M (52) | 220 | 444 | 2.02 (0.41, 9.92) | 0.05 |
| Malawi | iLiNS−DOSE (53) | | | | |
| **I² = 0.00, Tau² = 0.00** | | **5675** | **4365** | **0.86 (0.62, 1.20)** | |
| **Source water quality − Unimproved** | | | | | |
| Burkina Faso | iLiNS−Zinc (45) | 1952 | 664 | 0.36 (0.19, 0.68) | 0.29 |
| Burkina Faso | PROMIS (46) | 856 | 907 | 1.06 (0.33, 3.38) | 0.09 |
| Burkina Faso | PROMIS CS (46) | 430 | 436 | 0.51 (0.17, 1.54) | 0.10 |
| Kenya | WASH−B (50) | 694 | 2244 | 0.46 (0.07, 2.88) | 0.04 |
| Madagascar | MAHAY (51) | 1700 | 1682 | 0.58 (0.29, 1.17) | 0.24 |
| Mali | PROMIS (54) | | | | |
| Mali | PROMIS CS (54) | 944 | 959 | 0.48 (0.21, 1.10) | 0.18 |
| Zimbabwe | SHINE (HIV−) (55) | 887 | 876 | 0.79 (0.23, 2.76) | 0.08 |
| Zimbabwe | SHINE (HIV+) (56) | | | | |
| **I² = 0.00, Tau² = 0.00** | | **7463** | **7768** | **0.52 (0.37, 0.73)** | |

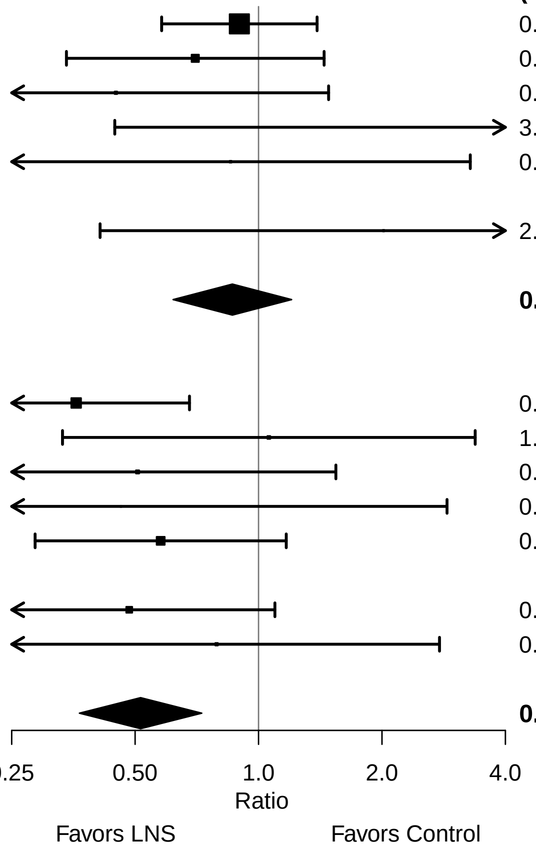

Supplemental figure 8: Severe wasting prevalence ratio

#### 8F: Stratified by Sanitation

**Sanitation**
**(p−diff = 0.154)**

| Country | Trial | N | N | PR (95% CI) | W |
|---|---|---|---|---|---|
| **Sanitation − Improved** | | | | | |
| Bangladesh | JiVitA−4 (42) | 2783 | 1217 | 0.90 (0.58, 1.39) | 0.44 |
| Bangladesh | RDNS (43) | 1661 | 815 | 0.70 (0.34, 1.45) | 0.17 |
| Bangladesh | WASH−B (44) | 566 | 1101 | 0.45 (0.14, 1.48) | 0.06 |
| Burkina Faso | PROMIS (46) | 856 | 907 | 1.06 (0.33, 3.38) | 0.07 |
| Burkina Faso | PROMIS CS (46) | 430 | 436 | 0.51 (0.17, 1.54) | 0.07 |
| Ghana | GHANA (47) | 98 | 96 | 3.92 (0.45, 34.42) | 0.02 |
| Ghana | iLiNS−DYAD−G (48) | 347 | 692 | 0.85 (0.22, 3.28) | 0.05 |
| Haiti | HAITI (49) | | | | |
| Mali | PROMIS (54) | | | | |
| Mali | PROMIS CS (54) | 944 | 959 | 0.48 (0.21, 1.10) | 0.13 |
| **I² = 0.00, Tau² = 0.00** | | **7685** | **6223** | **0.76 (0.57, 1.02)** | |
| **Sanitation − Unimproved** | | | | | |
| Burkina Faso | iLiNS−Zinc (45) | 1952 | 664 | 0.36 (0.19, 0.68) | 0.42 |
| Kenya | WASH−B (50) | 694 | 2244 | 0.46 (0.07, 2.88) | 0.05 |
| Madagascar | MAHAY (51) | 1700 | 1682 | 0.58 (0.29, 1.17) | 0.35 |
| Malawi | iLiNS−DYAD−M (52) | 220 | 444 | 2.02 (0.41, 9.92) | 0.07 |
| Malawi | iLiNS−DOSE (53) | | | | |
| Zimbabwe | SHINE (HIV−) (55) | 887 | 876 | 0.79 (0.23, 2.76) | 0.11 |
| Zimbabwe | SHINE (HIV+) (56) | | | | |
| **I² = 0.13, Tau² = 0.04** | | **5453** | **5910** | **0.54 (0.34, 0.87)** | |

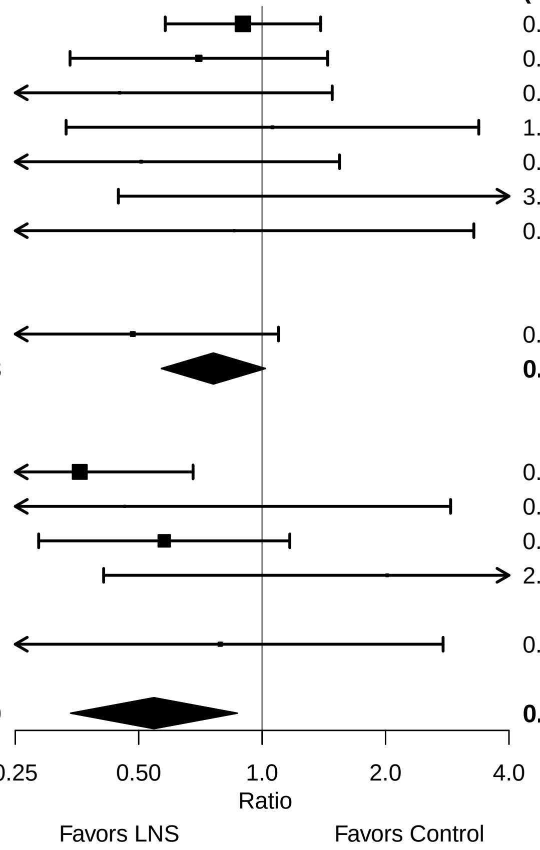

Supplemental figure 8: Severe wasting prevalence ratio

#### 8G: Stratified by Supplement duration

**Supplement duration**
**(p−diff = 0.375)**

**Supplement duration − 12m or less**

| **Country** | **Trial** | **N** | **N** | **PR (95% CI)** | **W** |
|---|---|---|---|---|---|
| Bangladesh | JiVitA−4 (42) | 2783 | 1217 | 0.90 (0.58, 1.39) | 0.37 |
| Burkina Faso | iLiNS−Zinc (45) | 1952 | 664 | 0.36 (0.19, 0.68) | 0.18 |
| Burkina Faso | PROMIS (46) | 856 | 907 | 1.06 (0.33, 3.38) | 0.05 |
| Burkina Faso | PROMIS CS (46) | 430 | 436 | 0.51 (0.17, 1.54) | 0.06 |
| Ghana | GHANA (47) | 98 | 96 | 3.92 (0.45, 34.42) | 0.02 |
| Ghana | iLiNS−DYAD−G (48) | 347 | 692 | 0.85 (0.22, 3.28) | 0.04 |
| Haiti | HAITI (49) | | | | |
| Madagascar | MAHAY (51) | 1700 | 1682 | 0.58 (0.29, 1.17) | 0.15 |
| Malawi | iLiNS−DYAD−M (52) | 220 | 444 | 2.02 (0.41, 9.92) | 0.03 |
| Malawi | iLiNS−DOSE (53) | | | | |
| Zimbabwe | SHINE (HIV−) (55) | 1869 | 1784 | 0.95 (0.38, 2.38) | 0.09 |
| Zimbabwe | SHINE (HIV+) (56) | 336 | 328 | 0.98 (0.14, 6.63) | 0.02 |
| **I² = 0.20, Tau² = 0.04** | | **10591** | **8250** | **0.74 (0.54, 1.02)** | |

**Supplement duration − > 12m**

| **Country** | **Trial** | **N** | **N** | **PR (95% CI)** | **W** |
|---|---|---|---|---|---|
| Bangladesh | RDNS (43) | 1661 | 815 | 0.70 (0.34, 1.45) | 0.34 |
| Bangladesh | WASH−B (44) | 1156 | 3424 | 0.61 (0.30, 1.25) | 0.34 |
| Kenya | WASH−B (50) | 1455 | 5118 | 0.27 (0.04, 1.86) | 0.05 |
| Mali | PROMIS (54) | | | | |
| Mali | PROMIS CS (54) | 944 | 959 | 0.48 (0.21, 1.10) | 0.27 |
| **I² = 0.00, Tau² = 0.00** | | **5216** | **10316** | **0.58 (0.38, 0.88)** | |

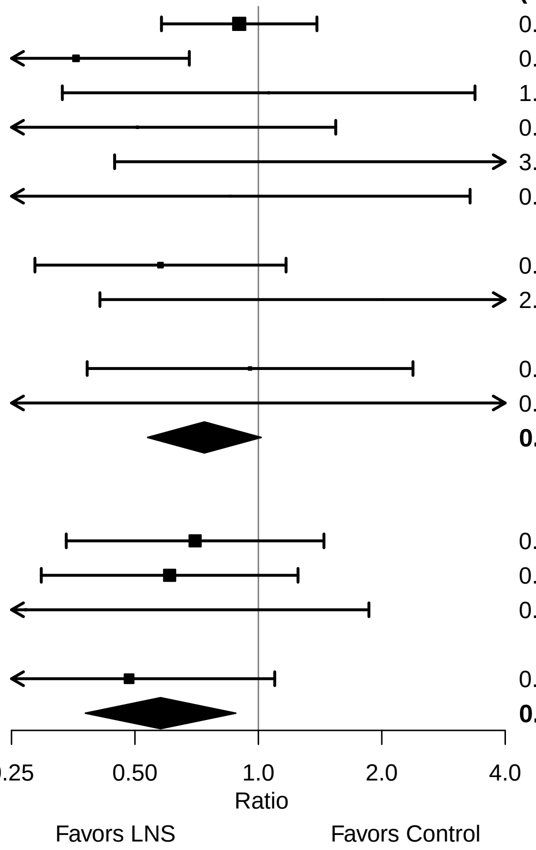

**Supplemental figure 8: Severe wasting prevalence ratio**

#### 8H: Stratified by Frequency of contact

**Frequency of contact**
**(p−diff = 0.710)**

| Country | Trial | N | N | PR (95% CI) | W |
|---|---|---|---|---|---|
| **Frequency of contact − Monthly** | | | | | |
| Bangladesh | RDNS (43) | 1661 | 815 | 0.70 (0.34, 1.45) | 0.22 |
| Burkina Faso | PROMIS (46) | 856 | 907 | 1.06 (0.33, 3.38) | 0.09 |
| Burkina Faso | PROMIS CS (46) | 430 | 436 | 0.51 (0.17, 1.54) | 0.09 |
| Haiti | HAITI (49) | | | | |
| Kenya | WASH−B (50) | 1455 | 5118 | 0.27 (0.04, 1.86) | 0.03 |
| Madagascar | MAHAY (51) | 1700 | 1682 | 0.58 (0.29, 1.17) | 0.23 |
| Mali | PROMIS (54) | | | | |
| Mali | PROMIS CS (54) | 944 | 959 | 0.48 (0.21, 1.10) | 0.17 |
| Zimbabwe | SHINE (HIV−) (55) | 1869 | 1784 | 0.95 (0.38, 2.38) | 0.14 |
| Zimbabwe | SHINE (HIV+) (56) | 336 | 328 | 0.98 (0.14, 6.63) | 0.03 |
| **$I^2$ = 0.00, $Tau^2$ = 0.00** | | **9251** | **12029** | **0.65 (0.46, 0.91)** | |
| **Frequency of contact − Weekly** | | | | | |
| Bangladesh | JiVitA−4 (42) | 2783 | 1217 | 0.90 (0.58, 1.39) | 0.48 |
| Bangladesh | WASH−B (44) | 1156 | 3424 | 0.61 (0.30, 1.25) | 0.18 |
| Burkina Faso | iLiNS−Zinc (45) | 1952 | 664 | 0.36 (0.19, 0.68) | 0.23 |
| Ghana | GHANA (47) | 98 | 96 | 3.92 (0.45, 34.42) | 0.02 |
| Ghana | iLiNS−DYAD−G (48) | 347 | 692 | 0.85 (0.22, 3.28) | 0.05 |
| Malawi | iLiNS−DYAD−M (52) | 220 | 444 | 2.02 (0.41, 9.92) | 0.04 |
| Malawi | iLiNS−DOSE (53) | | | | |
| **$I^2$ = 0.49, $Tau^2$ = 0.21** | | **6556** | **6537** | **0.76 (0.44, 1.31)** | |

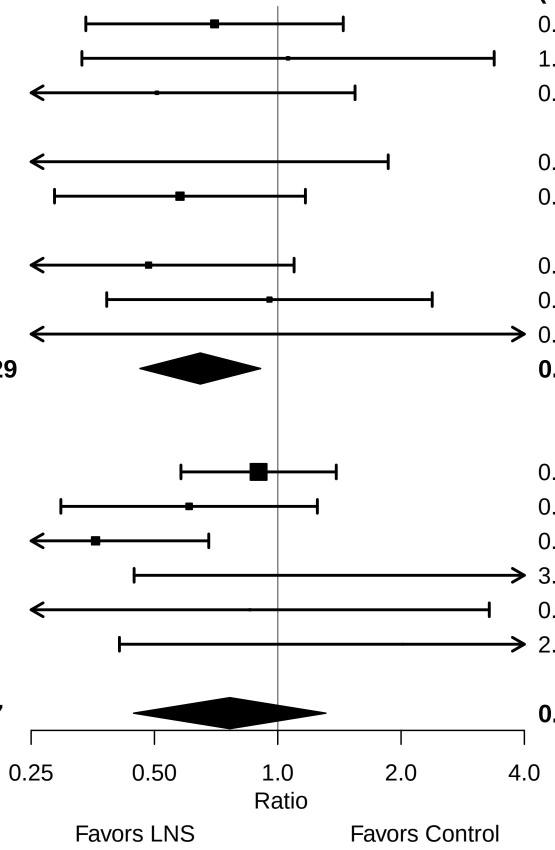

**Supplemental figure 8: Severe wasting prevalence ratio**

#### 8I: Stratified by Average SQ-LNS compliance

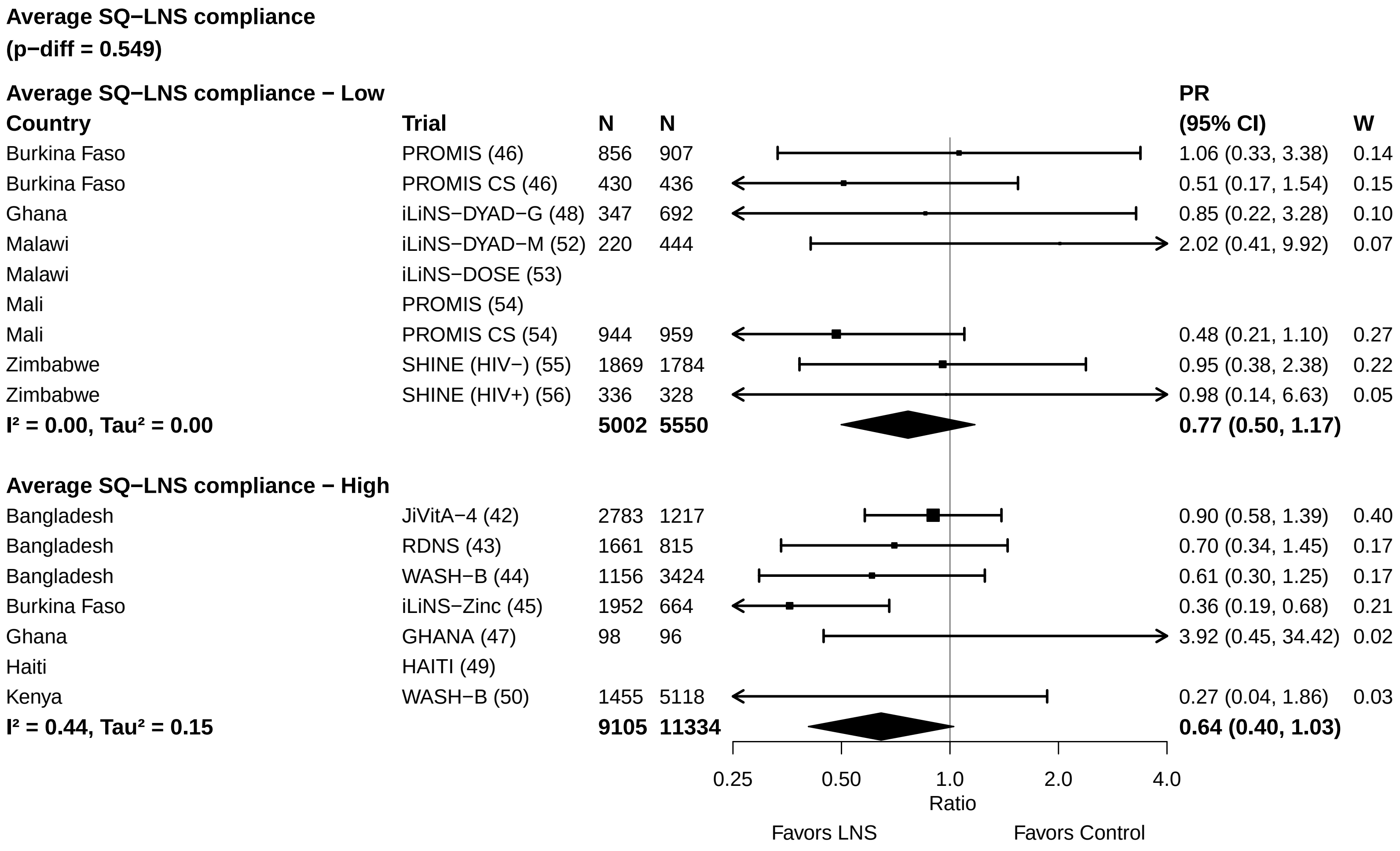
