## Supplemental Figure 9 for "Preventive small-quantity lipid-based nutrient supplements reduce severe wasting and severe stunting among young children: an individual participant data meta-analysis of randomized controlled trials"

### Supplemental figure 9: Forest plots for effects of SQ-LNS on severe stunting stratified by study-level effect modifiers

These figures are forest plots showing the study-level effect modification of intervention effects. Each figure shows the study-level estimates along with the corresponding pooled estimate grouped by study-level effect modifier category.
Individual study estimates were generated from log-binomial regression; controlling for baseline measure when available and with clustered observations using robust standard errors for cluster-randomized trials. Pooled sub-group estimates were generated using inverse-variance weighting random effects. P-value for the difference was estimated using random effects meta-regression with the indicated effect modifier as the predictor of intervention effect size; stratified pooled estimates are presented for each strata.

The labels on the left y-axis correspond to trial level information. The values on the right indicate the study level effect estimate, confidence interval, and weighting for deriving the pooled estimates.

### Supplemental figure 9: Severe stunting prevalence ratio

#### 9A: Stratified by Geographic region

**Geographic region**
**(p−diff = 0.970)**

| Country | Trial | N | N | PR (95% CI) | W |
|---|---|---|---|---|---|
| **Geographic region − SEAR** | | | | | |
| Bangladesh | JiVitA−4 (42) | 2838 | 1244 | 0.95 (0.78, 1.17) | 0.37 |
| Bangladesh | RDNS (43) | 1663 | 815 | 0.89 (0.67, 1.20) | 0.27 |
| Bangladesh | WASH−B (44) | 1158 | 3431 | 0.71 (0.58, 0.87) | 0.37 |
| **I² = 0.52, Tau² = 0.01** | | **5659** | **5490** | **0.84 (0.70, 1.01)** | |
| **Geographic region − AFR** | | | | | |
| Burkina Faso | iLiNS−Zinc (45) | 1952 | 664 | 0.48 (0.38, 0.60) | 0.13 |
| Burkina Faso | PROMIS (46) | 863 | 914 | 0.96 (0.63, 1.47) | 0.06 |
| Burkina Faso | PROMIS CS (46) | 430 | 439 | 0.88 (0.42, 1.82) | 0.03 |
| Ghana | GHANA (47) | 98 | 96 | 0.98 (0.06, 15.43) | 0.00 |
| Ghana | iLiNS−DYAD−G (48) | 347 | 692 | 1.71 (0.58, 5.05) | 0.01 |
| Kenya | WASH−B (50) | 1457 | 5137 | 0.82 (0.67, 1.01) | 0.14 |
| Madagascar | MAHAY (51) | 1702 | 1682 | 1.01 (0.85, 1.21) | 0.15 |
| Malawi | iLiNS−DYAD−M (52) | 220 | 444 | 0.90 (0.55, 1.47) | 0.05 |
| Malawi | iLiNS−DOSE (53) | 696 | 241 | 0.97 (0.70, 1.35) | 0.09 |
| Mali | PROMIS (54) | 506 | 506 | 0.97 (0.64, 1.47) | 0.07 |
| Mali | PROMIS CS (54) | 952 | 969 | 0.73 (0.51, 1.06) | 0.08 |
| Zimbabwe | SHINE (HIV−) (55) | 1880 | 1794 | 0.82 (0.65, 1.04) | 0.13 |
| Zimbabwe | SHINE (HIV+) (56) | 337 | 330 | 0.89 (0.60, 1.30) | 0.07 |
| **I² = 0.61, Tau² = 0.03** | | **11440** | **13908** | **0.83 (0.73, 0.96)** | |

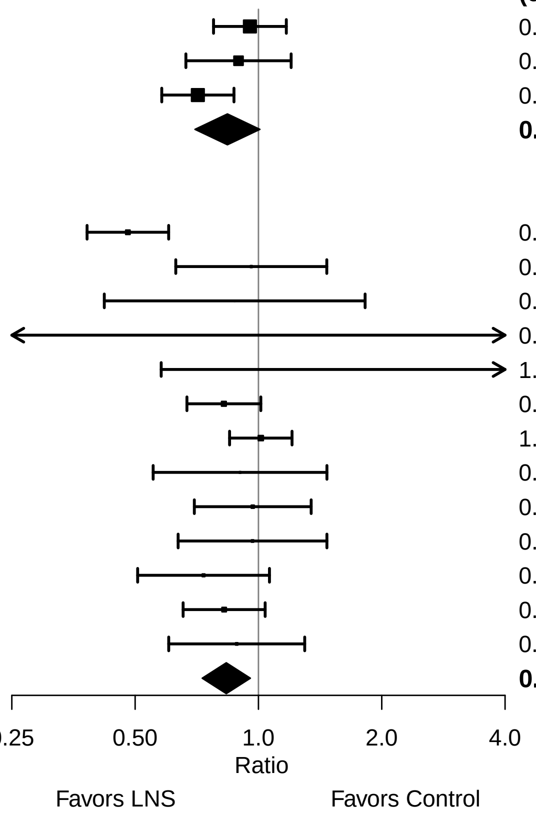

Supplemental figure 9: Severe stunting prevalence ratio

#### 9B: Stratified by Stunting burden

**Stunting burden**
**(p−diff = 0.309)**

| Country | Trial | N | N | PR (95% CI) | W |
|---|---|---|---|---|---|
| **Stunting burden − Less than 35%** | | | | | |
| Burkina Faso | PROMIS (46) | 863 | 914 | 0.96 (0.63, 1.47) | 0.21 |
| Burkina Faso | PROMIS CS (46) | 430 | 439 | 0.88 (0.42, 1.82) | 0.09 |
| Ghana | GHANA (47) | 98 | 96 | 0.98 (0.06, 15.43) | 0.01 |
| Ghana | iLiNS−DYAD−G (48) | 347 | 692 | 1.71 (0.58, 5.05) | 0.04 |
| Haiti | HAITI (49) | 149 | 149 | 1.79 (0.73, 4.42) | 0.06 |
| Kenya | WASH−B (50) | 1457 | 5137 | 0.82 (0.67, 1.01) | 0.42 |
| Malawi | iLiNS−DYAD−M (52) | 220 | 444 | 0.90 (0.55, 1.47) | 0.17 |
| **$I^2$ = 0.00, $Tau^2$ = 0.00** | | **3564** | **7871** | **0.89 (0.76, 1.05)** | |
| **Stunting burden − More than 35%** | | | | | |
| Bangladesh | JiVitA−4 (42) | 2838 | 1244 | 0.95 (0.78, 1.17) | 0.12 |
| Bangladesh | RDNS (43) | 1663 | 815 | 0.89 (0.67, 1.20) | 0.09 |
| Bangladesh | WASH−B (44) | 1158 | 3431 | 0.71 (0.58, 0.87) | 0.13 |
| Burkina Faso | iLiNS−Zinc (45) | 1952 | 664 | 0.48 (0.38, 0.60) | 0.12 |
| Madagascar | MAHAY (51) | 1702 | 1682 | 1.01 (0.85, 1.21) | 0.14 |
| Malawi | iLiNS−DOSE (53) | 696 | 241 | 0.97 (0.70, 1.35) | 0.08 |
| Mali | PROMIS (54) | 506 | 506 | 0.97 (0.64, 1.47) | 0.06 |
| Mali | PROMIS CS (54) | 952 | 969 | 0.73 (0.51, 1.06) | 0.07 |
| Zimbabwe | SHINE (HIV−) (55) | 1880 | 1794 | 0.82 (0.65, 1.04) | 0.12 |
| Zimbabwe | SHINE (HIV+) (56) | 337 | 330 | 0.89 (0.60, 1.30) | 0.07 |
| **$I^2$ = 0.73, $Tau^2$ = 0.04** | | **13684** | **11676** | **0.82 (0.71, 0.95)** | |

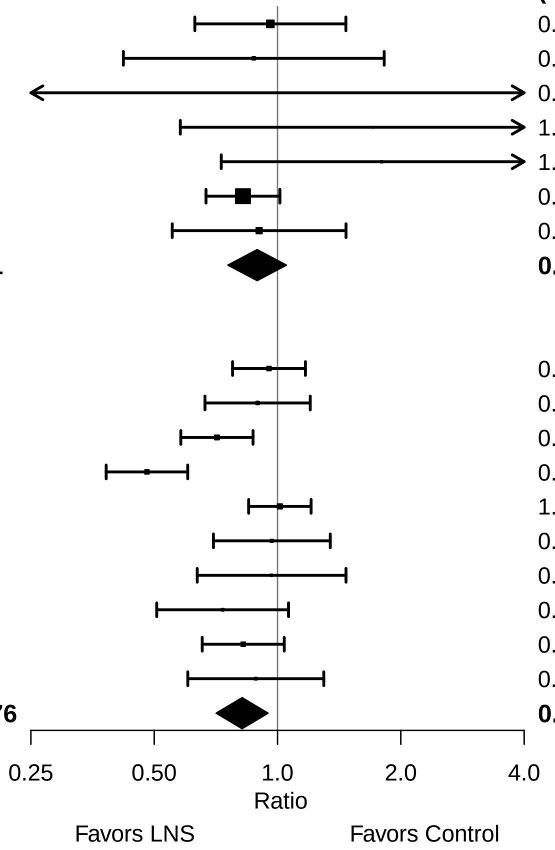

Supplemental figure 9: Severe stunting prevalence ratio

#### 9C: Stratified by Wasting burden

**Wasting burden**
**(p−diff = 0.123)**

**Wasting burden − Wasting < 10%**

| **Country** | **Trial** | **N** | **N** | **PR (95% CI)** | **W** |
|---|---|---|---|---|---|
| Ghana | GHANA (47) | 98 | 96 | 0.98 (0.06, 15.43) | 0.00 |
| Ghana | iLiNS−DYAD−G (48) | 347 | 692 | 1.71 (0.58, 5.05) | 0.02 |
| Haiti | HAITI (49) | 149 | 149 | 1.79 (0.73, 4.42) | 0.03 |
| Kenya | WASH−B (50) | 1457 | 5137 | 0.82 (0.67, 1.01) | 0.20 |
| Madagascar | MAHAY (51) | 1702 | 1682 | 1.01 (0.85, 1.21) | 0.22 |
| Malawi | iLiNS−DYAD−M (52) | 220 | 444 | 0.90 (0.55, 1.47) | 0.08 |
| Malawi | iLiNS−DOSE (53) | 696 | 241 | 0.97 (0.70, 1.35) | 0.14 |
| Zimbabwe | SHINE (HIV−) (55) | 1880 | 1794 | 0.82 (0.65, 1.04) | 0.19 |
| Zimbabwe | SHINE (HIV+) (56) | 337 | 330 | 0.89 (0.60, 1.30) | 0.11 |
| **$I^2$ = 0.00, $Tau^2$ = 0.00** | | **6886** | **10565** | **0.92 (0.83, 1.02)** | |

**Wasting burden − Wasting >= 10%**

| **Country** | **Trial** | **N** | **N** | **PR (95% CI)** | **W** |
|---|---|---|---|---|---|
| Bangladesh | JiVitA−4 (42) | 2838 | 1244 | 0.95 (0.78, 1.17) | 0.18 |
| Bangladesh | RDNS (43) | 1663 | 815 | 0.89 (0.67, 1.20) | 0.14 |
| Bangladesh | WASH−B (44) | 1158 | 3431 | 0.71 (0.58, 0.87) | 0.18 |
| Burkina Faso | iLiNS−Zinc (45) | 1952 | 664 | 0.48 (0.38, 0.60) | 0.17 |
| Burkina Faso | PROMIS (46) | 863 | 914 | 0.96 (0.63, 1.47) | 0.09 |
| Burkina Faso | PROMIS CS (46) | 430 | 439 | 0.88 (0.42, 1.82) | 0.04 |
| Mali | PROMIS (54) | 506 | 506 | 0.97 (0.64, 1.47) | 0.09 |
| Mali | PROMIS CS (54) | 952 | 969 | 0.73 (0.51, 1.06) | 0.11 |
| **$I^2$ = 0.71, $Tau^2$ = 0.04** | | **10362** | **8982** | **0.78 (0.65, 0.93)** | |

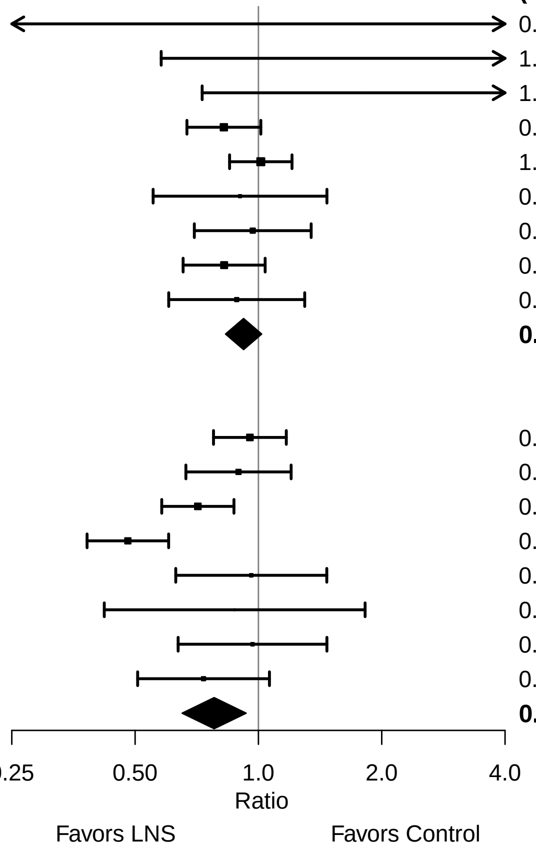

Supplemental figure 9: Severe stunting prevalence ratio

#### 9D: Stratified by Malaria prevalence

**Malaria prevalence**
**(p−diff = 0.407)**

**Malaria prevalence − Less than 10%**

| **Country** | **Trial** | **N** | **N** | **PR (95% CI)** | **W** |
|---|---|---|---|---|---|
| Bangladesh | JiVitA−4 (42) | 2838 | 1244 | 0.95 (0.78, 1.17) | 0.15 |
| Bangladesh | RDNS (43) | 1663 | 815 | 0.89 (0.67, 1.20) | 0.12 |
| Bangladesh | WASH−B (44) | 1158 | 3431 | 0.71 (0.58, 0.87) | 0.16 |
| Haiti | HAITI (49) | 149 | 149 | 1.79 (0.73, 4.42) | 0.02 |
| Kenya | WASH−B (50) | 1457 | 5137 | 0.82 (0.67, 1.01) | 0.15 |
| Madagascar | MAHAY (51) | 1702 | 1682 | 1.01 (0.85, 1.21) | 0.17 |
| Zimbabwe | SHINE (HIV−) (55) | 1880 | 1794 | 0.82 (0.65, 1.04) | 0.14 |
| Zimbabwe | SHINE (HIV+) (56) | 337 | 330 | 0.89 (0.60, 1.30) | 0.09 |
| **I² = 0.33, Tau² = 0.01** | | **11184** | **14582** | **0.88 (0.79, 0.98)** | |

**Malaria prevalence − At least 10%**

| **Country** | **Trial** | **N** | **N** | **PR (95% CI)** | **W** |
|---|---|---|---|---|---|
| Burkina Faso | iLiNS−Zinc (45) | 1952 | 664 | 0.48 (0.38, 0.60) | 0.24 |
| Burkina Faso | PROMIS (46) | 863 | 914 | 0.96 (0.63, 1.47) | 0.13 |
| Burkina Faso | PROMIS CS (46) | 430 | 439 | 0.88 (0.42, 1.82) | 0.05 |
| Ghana | GHANA (47) | 98 | 96 | 0.98 (0.06, 15.43) | 0.00 |
| Ghana | iLiNS−DYAD−G (48) | 347 | 692 | 1.71 (0.58, 5.05) | 0.03 |
| Malawi | iLiNS−DYAD−M (52) | 220 | 444 | 0.90 (0.55, 1.47) | 0.10 |
| Malawi | iLiNS−DOSE (53) | 696 | 241 | 0.97 (0.70, 1.35) | 0.17 |
| Mali | PROMIS (54) | 506 | 506 | 0.97 (0.64, 1.47) | 0.13 |
| Mali | PROMIS CS (54) | 952 | 969 | 0.73 (0.51, 1.06) | 0.15 |
| **I² = 0.64, Tau² = 0.04** | | **6064** | **4965** | **0.81 (0.65, 0.99)** | |

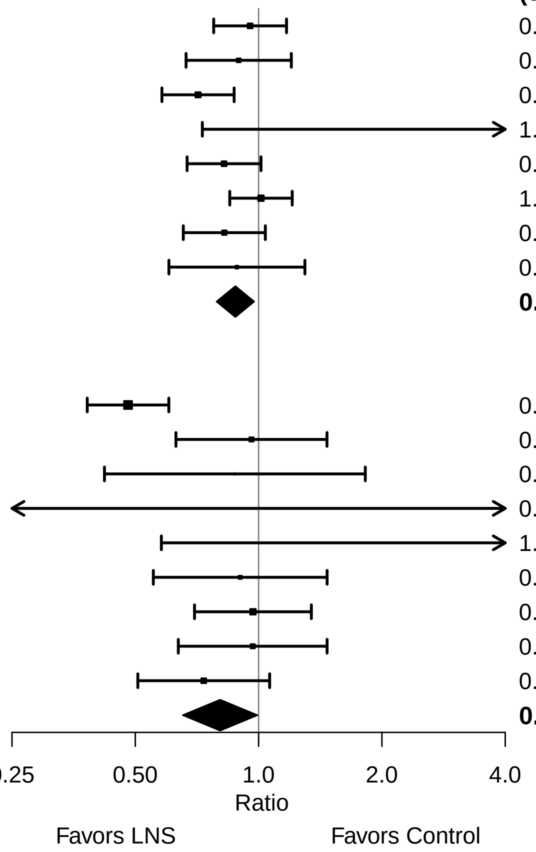

Supplemental figure 9: Severe stunting prevalence ratio

#### 9E: Stratified by Source water quality

**Source water quality**
**(p−diff = 0.369)**

**Source water quality − Improved**

| **Country** | **Trial** | **N** | **N** | **PR (95% CI)** | **W** |
|---|---|---|---|---|---|
| Bangladesh | JiVitA−4 (42) | 2838 | 1244 | 0.95 (0.78, 1.17) | 0.26 |
| Bangladesh | RDNS (43) | 1663 | 815 | 0.89 (0.67, 1.20) | 0.20 |
| Bangladesh | WASH−B (44) | 567 | 1104 | 0.73 (0.54, 1.00) | 0.19 |
| Ghana | GHANA (47) | 98 | 96 | 0.98 (0.06, 15.43) | 0.00 |
| Ghana | iLiNS−DYAD−G (48) | 347 | 692 | 1.71 (0.58, 5.05) | 0.03 |
| Haiti | HAITI (49) | 149 | 149 | 1.79 (0.73, 4.42) | 0.04 |
| Malawi | iLiNS−DYAD−M (52) | 220 | 444 | 0.90 (0.55, 1.47) | 0.11 |
| Malawi | iLiNS−DOSE (53) | 696 | 241 | 0.97 (0.70, 1.35) | 0.18 |
| **I² = 0.00, Tau² = 0.00** | | **6578** | **4785** | **0.92 (0.81, 1.04)** | |

**Source water quality − Unimproved**

| **Country** | **Trial** | **N** | **N** | **PR (95% CI)** | **W** |
|---|---|---|---|---|---|
| Burkina Faso | iLiNS−Zinc (45) | 1952 | 664 | 0.48 (0.38, 0.60) | 0.17 |
| Burkina Faso | PROMIS (46) | 863 | 914 | 0.96 (0.63, 1.47) | 0.09 |
| Burkina Faso | PROMIS CS (46) | 430 | 439 | 0.88 (0.42, 1.82) | 0.04 |
| Kenya | WASH−B (50) | 695 | 2253 | 0.87 (0.64, 1.19) | 0.13 |
| Madagascar | MAHAY (51) | 1702 | 1682 | 1.01 (0.85, 1.21) | 0.20 |
| Mali | PROMIS (54) | 506 | 506 | 0.97 (0.64, 1.47) | 0.09 |
| Mali | PROMIS CS (54) | 952 | 969 | 0.73 (0.51, 1.06) | 0.11 |
| Zimbabwe | SHINE (HIV−) (55) | 892 | 880 | 0.96 (0.68, 1.35) | 0.12 |
| Zimbabwe | SHINE (HIV+) (56) | 147 | 146 | 0.91 (0.54, 1.54) | 0.07 |
| **I² = 0.73, Tau² = 0.04** | | **8139** | **8453** | **0.83 (0.69, 0.99)** | |

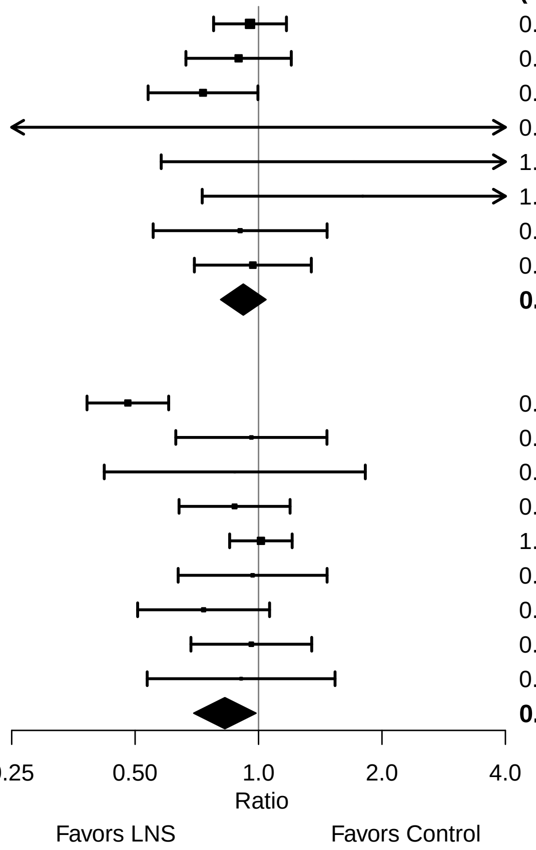

**Supplemental figure 9: Severe stunting prevalence ratio**

#### 9F: Stratified by Sanitation

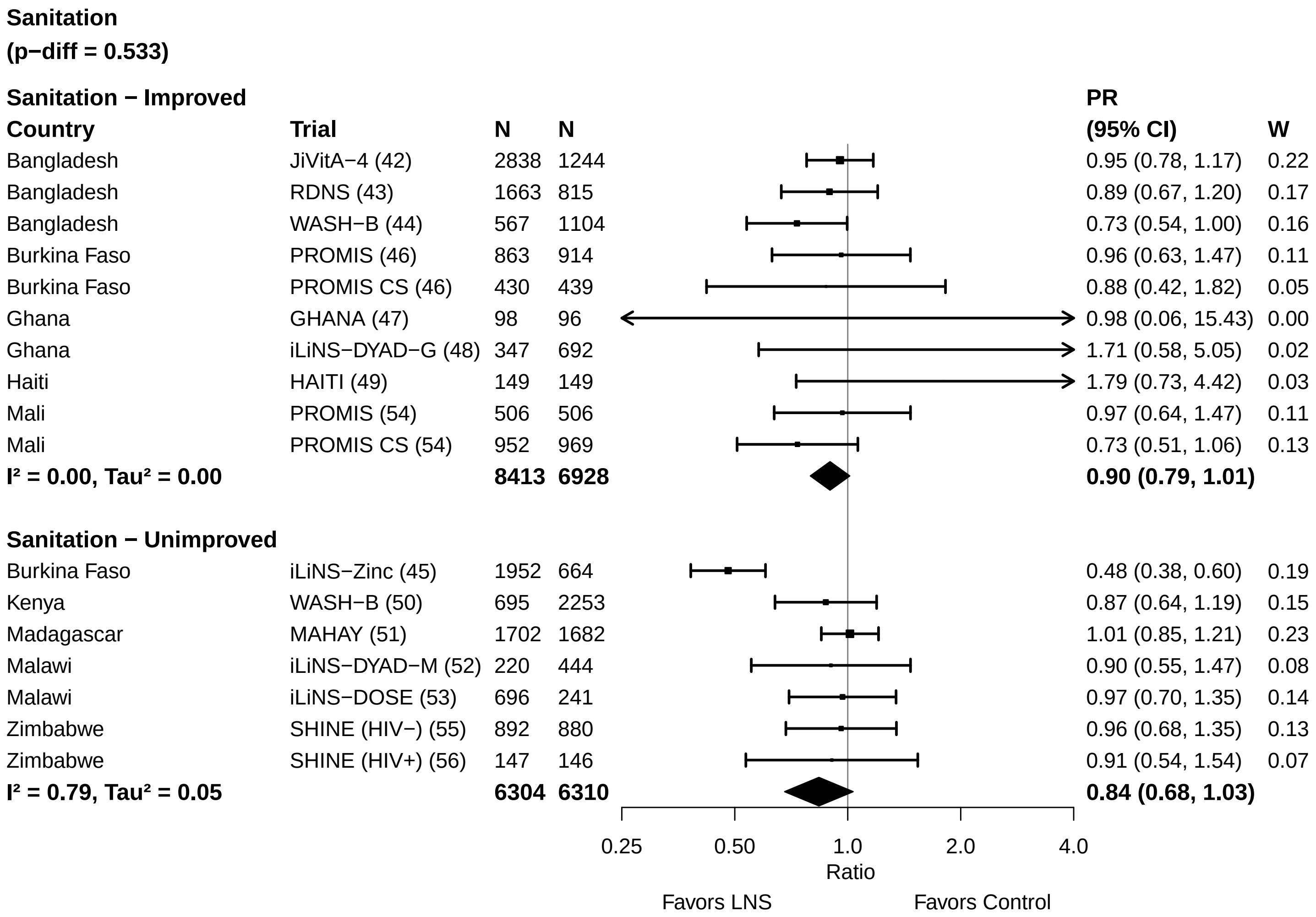

Supplemental figure 9: Severe stunting prevalence ratio

#### 9G: Stratified by Supplement duration

**Supplement duration**
**(p−diff = 0.572)**

| Country | Trial | N | N | PR (95% CI) | W |
|---|---|---|---|---|---|
| **Supplement duration − 12m or less** | | | | | |
| Bangladesh | JiVitA−4 (42) | 2838 | 1244 | 0.95 (0.78, 1.17) | 0.15 |
| Burkina Faso | iLiNS−Zinc (45) | 1952 | 664 | 0.48 (0.38, 0.60) | 0.14 |
| Burkina Faso | PROMIS (46) | 863 | 914 | 0.96 (0.63, 1.47) | 0.08 |
| Burkina Faso | PROMIS CS (46) | 430 | 439 | 0.88 (0.42, 1.82) | 0.03 |
| Ghana | GHANA (47) | 98 | 96 | 0.98 (0.06, 15.43) | 0.00 |
| Ghana | iLiNS−DYAD−G (48) | 347 | 692 | 1.71 (0.58, 5.05) | 0.02 |
| Haiti | HAITI (49) | 149 | 149 | 1.79 (0.73, 4.42) | 0.02 |
| Madagascar | MAHAY (51) | 1702 | 1682 | 1.01 (0.85, 1.21) | 0.17 |
| Malawi | iLiNS−DYAD−M (52) | 220 | 444 | 0.90 (0.55, 1.47) | 0.06 |
| Malawi | iLiNS−DOSE (53) | 696 | 241 | 0.97 (0.70, 1.35) | 0.10 |
| Zimbabwe | SHINE (HIV−) (55) | 1880 | 1794 | 0.82 (0.65, 1.04) | 0.14 |
| Zimbabwe | SHINE (HIV+) (56) | 337 | 330 | 0.89 (0.60, 1.30) | 0.09 |
| **I² = 0.68, Tau² = 0.04** | | **11512** | **8689** | **0.88 (0.74, 1.04)** | |
| **Supplement duration − > 12m** | | | | | |
| Bangladesh | RDNS (43) | 1663 | 815 | 0.89 (0.67, 1.20) | 0.19 |
| Bangladesh | WASH−B (44) | 1158 | 3431 | 0.71 (0.58, 0.87) | 0.26 |
| Kenya | WASH−B (50) | 1457 | 5137 | 0.82 (0.67, 1.01) | 0.26 |
| Mali | PROMIS (54) | 506 | 506 | 0.97 (0.64, 1.47) | 0.13 |
| Mali | PROMIS CS (54) | 952 | 969 | 0.73 (0.51, 1.06) | 0.15 |
| **I² = 0.00, Tau² = 0.00** | | **5736** | **10858** | **0.80 (0.71, 0.89)** | |

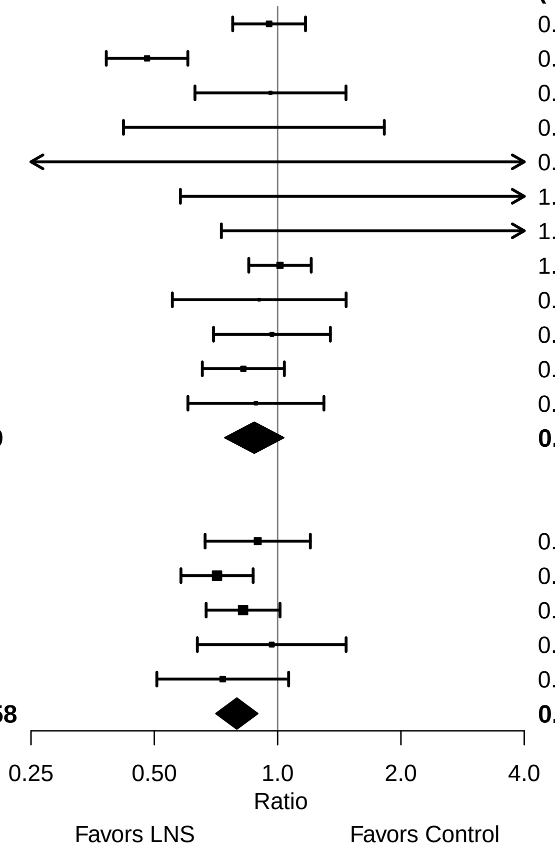

**Supplemental figure 9: Severe stunting prevalence ratio**

#### 9H: Stratified by Frequency of contact

**Frequency of contact**
**(p−diff = 0.220)**

**Frequency of contact − Monthly**

| **Country** | **Trial** | **N** | **N** | **PR (95% CI)** | **W** |
|---|---|---|---|---|---|
| Bangladesh | RDNS (43) | 1663 | 815 | 0.89 (0.67, 1.20) | 0.12 |
| Burkina Faso | PROMIS (46) | 863 | 914 | 0.96 (0.63, 1.47) | 0.08 |
| Burkina Faso | PROMIS CS (46) | 430 | 439 | 0.88 (0.42, 1.82) | 0.03 |
| Haiti | HAITI (49) | 149 | 149 | 1.79 (0.73, 4.42) | 0.02 |
| Kenya | WASH−B (50) | 1457 | 5137 | 0.82 (0.67, 1.01) | 0.16 |
| Madagascar | MAHAY (51) | 1702 | 1682 | 1.01 (0.85, 1.21) | 0.17 |
| Mali | PROMIS (54) | 506 | 506 | 0.97 (0.64, 1.47) | 0.08 |
| Mali | PROMIS CS (54) | 952 | 969 | 0.73 (0.51, 1.06) | 0.09 |
| Zimbabwe | SHINE (HIV−) (55) | 1880 | 1794 | 0.82 (0.65, 1.04) | 0.15 |
| Zimbabwe | SHINE (HIV+) (56) | 337 | 330 | 0.89 (0.60, 1.30) | 0.09 |
| **I² = 0.00, Tau² = 0.00** | | **9939** | **12735** | **0.90 (0.82, 0.99)** | |

**Frequency of contact − Weekly**

| **Country** | **Trial** | **N** | **N** | **PR (95% CI)** | **W** |
|---|---|---|---|---|---|
| Bangladesh | JiVitA−4 (42) | 2838 | 1244 | 0.95 (0.78, 1.17) | 0.24 |
| Bangladesh | WASH−B (44) | 1158 | 3431 | 0.71 (0.58, 0.87) | 0.24 |
| Burkina Faso | iLiNS−Zinc (45) | 1952 | 664 | 0.48 (0.38, 0.60) | 0.22 |
| Ghana | GHANA (47) | 98 | 96 | 0.98 (0.06, 15.43) | 0.00 |
| Ghana | iLiNS−DYAD−G (48) | 347 | 692 | 1.71 (0.58, 5.05) | 0.03 |
| Malawi | iLiNS−DYAD−M (52) | 220 | 444 | 0.90 (0.55, 1.47) | 0.10 |
| Malawi | iLiNS−DOSE (53) | 696 | 241 | 0.97 (0.70, 1.35) | 0.16 |
| **I² = 0.76, Tau² = 0.06** | | **7309** | **6812** | **0.79 (0.62, 1.02)** | |

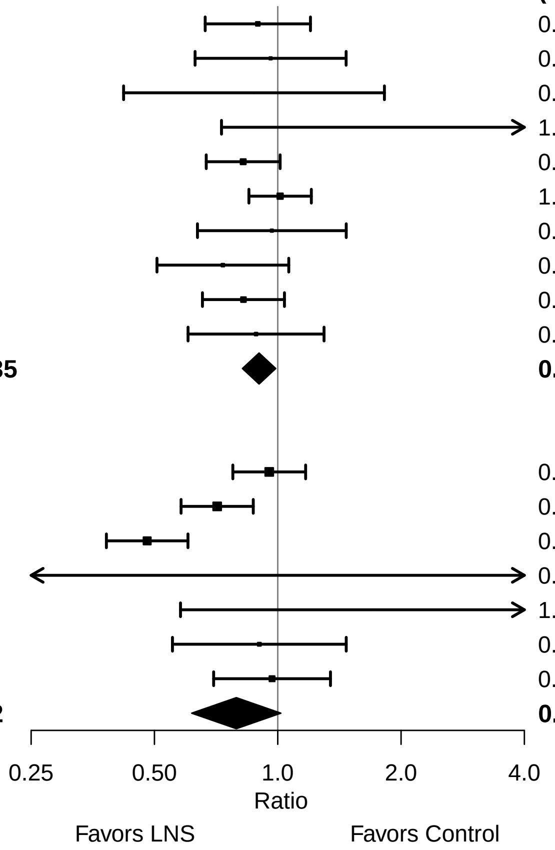

**Supplemental figure 9: Severe stunting prevalence ratio**

#### 9I: Stratified by Average SQ-LNS compliance

**Average SQ−LNS compliance**
**(p−diff = 0.251)**

**Average SQ−LNS compliance − Low**

| Country | Trial | N | N | PR (95% CI) | W |
|---|---|---|---|---|---|
| Burkina Faso | PROMIS (46) | 863 | 914 | 0.96 (0.63, 1.47) | 0.11 |
| Burkina Faso | PROMIS CS (46) | 430 | 439 | 0.88 (0.42, 1.82) | 0.05 |
| Ghana | iLiNS−DYAD−G (48) | 347 | 692 | 1.71 (0.58, 5.05) | 0.02 |
| Malawi | iLiNS−DYAD−M (52) | 220 | 444 | 0.90 (0.55, 1.47) | 0.09 |
| Malawi | iLiNS−DOSE (53) | 696 | 241 | 0.97 (0.70, 1.35) | 0.15 |
| Mali | PROMIS (54) | 506 | 506 | 0.97 (0.64, 1.47) | 0.11 |
| Mali | PROMIS CS (54) | 952 | 969 | 0.73 (0.51, 1.06) | 0.13 |
| Zimbabwe | SHINE (HIV−) (55) | 1880 | 1794 | 0.82 (0.65, 1.04) | 0.21 |
| Zimbabwe | SHINE (HIV+) (56) | 337 | 330 | 0.89 (0.60, 1.30) | 0.13 |
| **I² = 0.00, Tau² = 0.00** | | **6231** | **6329** | **0.88 (0.77, 1.00)** | |

**Average SQ−LNS compliance − High**

| Country | Trial | N | N | PR (95% CI) | W |
|---|---|---|---|---|---|
| Bangladesh | JiVitA−4 (42) | 2838 | 1244 | 0.95 (0.78, 1.17) | 0.21 |
| Bangladesh | RDNS (43) | 1663 | 815 | 0.89 (0.67, 1.20) | 0.15 |
| Bangladesh | WASH−B (44) | 1158 | 3431 | 0.71 (0.58, 0.87) | 0.21 |
| Burkina Faso | iLiNS−Zinc (45) | 1952 | 664 | 0.48 (0.38, 0.60) | 0.19 |
| Ghana | GHANA (47) | 98 | 96 | 0.98 (0.06, 15.43) | 0.00 |
| Haiti | HAITI (49) | 149 | 149 | 1.79 (0.73, 4.42) | 0.03 |
| Kenya | WASH−B (50) | 1457 | 5137 | 0.82 (0.67, 1.01) | 0.21 |
| **I² = 0.77, Tau² = 0.07** | | **9315** | **11536** | **0.79 (0.62, 1.01)** | |

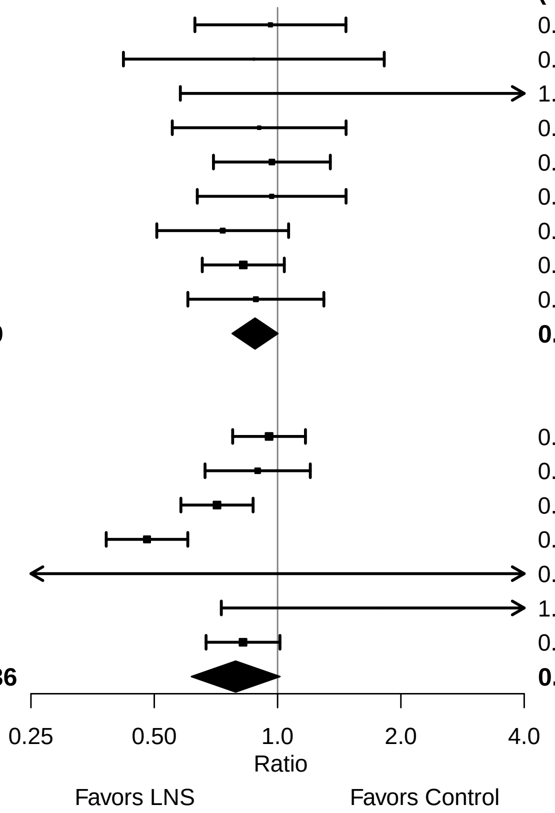
